# The genetic, clinical and immune landscape of clonal hematopoiesis across adulthood in the multi-ancestry Mount Sinai Million cohort

**DOI:** 10.64898/2026.10.22.26366221

**Authors:** Myvizhi Esai Selvan, Namira Ashraf, Rujula Pradeep, Leyla Giordano, Max Dougherty, Paul Chevelev, Carl Jannes Neuse, Daniel I. Nathan, Bridget K. Marcellino, Robert J. Klein, Zeynep H. Gümüş

## Abstract

Clonal hematopoiesis of indeterminate potential (CHIP), the age-associated expansion of blood cell clones carrying acquired driver mutations, remains less well characterized in young adults, across genetic ancestries, and in clinical populations. We analyzed whole-exome sequencing and longitudinal electronic health record (EHR) data from 59,598 adults in the Mount Sinai Million Health Discoveries Program, an urban, multi-ancestry hospital-based cohort that includes substantial representation of African (∼35%) and Admixed American genetic ancestry (∼15%) and young adults aged 18-39 years (∼30%). After sample QC, separate analyses were performed in discovery (n = 28,051) and validation (n = 26,938) cohorts defined by sequencing platform and combined by meta-analysis.

CHIP was detected in 8.5% of participants in each cohort and in 4.0% of young adults, with D*NMT3A, TET2, ASXL1,* and *PPM1D* the most frequently mutated genes. Prevalence and clone size increased with age across ancestry groups. After adjusting for age, participants of African ancestry had lower CHIP prevalence than those of European ancestry in both cohorts (discovery: OR=0.86, 95% CI = 0.78–0.95, *p*=0.002; validation: OR=0.84, 95% CI = 0.75–0.93, *p*=0.001). Participants of Admixed American ancestry showed a similar trend.

Higher systemic immune-inflammation index (SII) was associated with CHIP after adjusting for age, sex and ancestry in both cohorts (pooled OR=1.30, 95% CI = 1.15–1.48, *p*=4.78e-5). Heterozygosity at an HLA class II locus *HLA-DPA1* was associated with 7% lower odds of overall CHIP (pooled OR=0.93, 95% CI = 0.87–0.99, *p*=0.03). Driver-stratified analyses revealed distinct HLA class II nominal association patterns for *DNMT3A*- and *TET2*-mutant CHIP. Particularly, *HLA-DQB1* heterozygosity associated with lower odds of *TET2*-mutant CHIP (pooled OR=0.84, 95% CI = 0.70–1.00, *p*=0.05), *HLA-DQA1* and *HLA-DOB* heterozygosity associated with higher odds of *DNMT3A*-mutant CHIP (pooled OR=1.19, 95% CI = 1.00–1.42, *p*=0.05; pooled OR=1.12, 95% CI = 1.01–1.25, *p*=0.03 respectively). Longitudinal EHR analyses revealed CHIP associations with several incident diseases that varied by driver gene and clone size. The broadest pattern of associations involved high-risk CHIP genes and encompassed hematologic malignancies, solid tumors, cardiovascular outcomes, hypertension, and COPD. High-VAF clones were the main drivers of these associations. Among adults aged ≥40 years, phenome-wide analyses identified a Bonferroni-significant association with incident myeloproliferative disease and nominal associations with incident pancytopenia, anemia, cardiomegaly, and chronic renal failure. Young-adult CHIP was nominally associated with incident nodular goiter and esophageal disease. These findings characterize CHIP across adulthood and genetic ancestry and reveal inherited immune variation and systemic inflammation as correlates in a hospital-based population.

## Introduction

Clonal hematopoiesis of indeterminate potential (CHIP) is an aging-associated expansion of hematopoietic stem and progenitor cell clones carrying acquired leukemogenic mutations without clinical evidence of a hematologic malignancy^1^. The World Health Organization (WHO) defines CHIP using a variant allele fraction (VAF) threshold of at least 2%^2^. CHIP is uncommon in early adulthood but affects 10-20% of adults over 70 years^3^. Most carriers harbor a single mutation, commonly in the epigenetic regulators *DNMT3A*, *TET2*, and *ASXL1*. CHIP has been associated with a 10- to 13-fold increased risk of hematologic malignancy, a nearly 2-fold increased risk of coronary heart disease and elevated all-cause mortality^1,4–6^. These risks vary according to the underlying driver mutation and clone size.

Large sequencing studies, including the UK Biobank, Trans-Omics for Precision Medicine (TOPMed), and the All of Us Research Program have substantially expanded our understanding of the genetic and clinical landscape of CHIP^7–9^. However, estimates of CHIP prevalence, driver composition, and clinical associations may vary with cohort ascertainment, age distribution, genetic ancestry, and sequencing technology^7,9,10^. Younger adults remain underrepresented in biobank-scale sequencing studies, which are weighted toward middle-aged and older participants. This limited representation, together with the low prevalence of CHIP detectable by standard biobank sequencing before age 40, leaves its mutational spectrum and clinical correlates in younger adults comparatively poorly characterized^4,9^.

Beyond differences in population representation, inherited genetic variation may contribute to heterogeneity in CHIP. Although its defining mutations are acquired, population-scale studies have identified common and rare germline susceptibility variants, including associations with specific CHIP drivers^7,11^. Inherited variation in the highly polymorphic human leukocyte antigen (HLA) region may influence immune recognition of mutant clones through differences in antigen presentation. HLA heterozygosity may broaden the repertoire of presented peptides; class I heterozygosity has been associated with improved outcomes following immune-checkpoint blockade^12^ and class II heterozygosity with reduced lung cancer risk among smokers^13^. While recent UK Biobank study found no consistent association between predicted binding of mutant peptides to HLA molecules and mutation prevalence or clone size^14^, it only focused on 40 recurrent hotspot variants, did not include *TET2*, and evaluated only classical HLA loci.

Alongside potential differences in immune recognition, systemic inflammation may impact the expansion and biological effects of mutant clones. Experimental studies support a bidirectional relationship in which CHIP-associated mutations alter inflammatory signaling, while inflammatory stress can confer a selective advantage to mutant cells^15–17^. For example, *TET2* deficiency enhances *NLRP3* inflammasome–dependent *IL-1β* signaling, whereas inflammatory stress promotes the survival and expansion of *TET2*-mutant clones^15,16^. These effects may vary by driver gene^17^. Composite indices derived from routine laboratory measurements offer a scalable approach to assessing the inflammatory correlates of CHIP. The systemic immune-inflammation index (SII), integrates neutrophil, lymphocyte and platelet counts, whereas the INFLA-score combines C-reactive protein with cellular measures of low-grade inflammation^18–20^. Large hospital-based cohorts linked to longitudinal electronic health records (EHRs) enable the examination of CHIP across a broad age range, multiple genetic ancestry groups, and a wide spectrum of clinical conditions. These data also support phenome-wide association studies (PheWAS) to characterize clinical correlates across adulthood. As both CHIP prevalence and disease burden increase strongly with age, age-stratified analyses may identify clinical associations that would be obscured in a pooled population.

Here, we analyzed whole-exome sequencing (WES) and longitudinal EHR data from ∼60,000 adults in the Mount Sinai Million (MSM) Health Discoveries Program, an urban hospital-based cohort with substantial representation of African (∼35%) and Admixed American genetic ancestry (∼15%) and young adults aged 18–39 years (∼30%). Participants were divided into discovery and validation cohorts to account for differences in sequencing platforms. We characterized the CHIP mutational landscape across age and genetic ancestry groups, including a focused analysis of adults aged 18-39 years; examined inherited susceptibility, with particular emphasis on HLA class I and class II heterozygosity; evaluated associations with systemic immune-inflammation index SII; and performed outcome-specific association analyses and age-stratified PheWAS. Together, these analyses characterize the demographic, immune, and clinical heterogeneity of CHIP across adulthood in an urban, multi-ancestry, EHR-linked hospital population.

## Methods

### Study design and population

The Mount Sinai Million (MSM) Health Discoveries Program (https://mountsinaimillion.org) is a health system-wide precision medicine initiative designed to enroll up to one million patients receiving care across the Mount Sinai Health System (MSHS). MSM builds on the BioMe Biobank^21^, an electronic health record (EHR)-linked biobank established in 2006 whose enrollment concluded in 2022. Both programs link genomic and biospecimen data with longitudinal EHR-derived clinical information. As MSHS serves diverse communities throughout New York City, the combined resource includes participants spanning a broad range of genetic ancestries and clinical conditions. At the time of this analysis, approximately 300,000 participants were enrolled, and genomic data were available for 59,598 participants.

The discovery cohort comprised 31,250 participants enrolled through BioMe before 2022. Whole-exome sequencing (WES) was performed using the IDT xGen capture panel (Integrated DNA Technologies, Inc, Coralville, IA) and an Illumina v4 HiSeq 2500 platform. Genome-wide genotyping was performed using the Infinium Global Screening Array (GSA). The validation cohort comprised 28,348 participants enrolled through MSM beginning in 2022, and genotyped using both genotype-by-sequencing (GxS) and a Twist targeted sequencing panel (Twist Bioscience, San Francisco, CA). Analyses were conducted separately in the discovery and validation cohorts to account for differences in the genomic platforms and sequencing depth, with cohort-specific results combined using meta-analysis where specified. The mean depth of coverage is ∼36x^22^ for the discovery cohort and ∼45x for the validation cohort which could lead to differences in variant detection and CHIP prevalence between cohorts. Participants in the discovery cohort were enrolled earlier, with a median follow-up of 8.9 years, versus the validation cohort at 3.8 years. Follow-up time was defined as the time interval, in years, from specimen collection to the latest available diagnosis encounter for each participant.

The Program for the Protection of Human Subjects (PPHS) at the Icahn School of Medicine at Mount Sinai determined that this study met the criteria for exempt human-subjects research (**IRB no.** 20-03271).

### Participant selection and sample quality control (QC)

Of the 59,598 participants with WES data, 587 were excluded during sample-level quality control (QC) due to discordance between genetically inferred and reported sex, estimated contamination >5%, <80% of targeted exome bases achieving >20× coverage, or unresolved duplicates. After sample QC, 59,011 participants remained (30,784 in the discovery cohort and 28,227 in the validation cohort). We then excluded participants with missing phenotype or EHR data, duplicate or conflicting phenotype records, or age <18 years at biospecimen collection. We also excluded participants with a diagnosis of a lymphoid, hematopoietic, or related tissue neoplasm recorded before or within six months after biospecimen collection (ICD-10: C81-C96, D45-D47, Z85.6 and Z85.7; ICD-9: 200-208, 238.4-238.7, V10.6 and V10.7). This six-month window was used to reduce the likelihood that variants arising from an existing or recently diagnosed hematologic malignancy would be classified as clonal hematopoiesis. The final study population comprised 54,989 participants: 28,051 in the discovery cohort and 26,938 in the validation cohort. Participant demographic and clinical characteristics are summarized in Table 1. Note that participants with baseline cytopenias were included in the primary analysis. Clonal cytopenia of undetermined significance (CCUS) was not clinically adjudicated, as no dedicated ICD-9-CM/ICD-10-CM code was available for its identification.

**Table 1.** Discovery and validation cohort characteristics.

| **Phenotype** | **Category** | **Discovery cohort (28,051)**<br>**# Samples (%)** | **Validation cohort (26,938)**<br>**# Samples (%)** |
| --- | --- | --- | --- |
| Sex | Female | 16,687 (59.5) | 15,705 (58.3) |
|  | Male | 11,364 (40.5) | 11,233 (41.7) |
| Smoking status | No | 22,700 (80.9) | 11,298 (41.9) |
|  | Yes | 4,954 (17.7) | 2,959 (11.0) |
|  | Missing | 397 (1.4) | 12,681 (47.1) |
| Genetic ancestry | EUR | 9,994 (35.6) | 10,240 (38.0) |
|  | AFR | 11,518 (41.1) | 7,494 (27.8) |
|  | AMR | 3,267 (11.6) | 4,920 (18.3) |
|  | EAS | 792 (2.8) | 1,576 (5.9) |
|  | SAS | 801 (2.9) | 1,182 (4.4) |
|  | Unknown | 1,679 (6.0) | 1,526 (5.7) |
| Age | < 40 | 7,337 (26.2) | 8,637 (32.1) |
|  | ≥ 40 and <50 | 5,100 (18.2) | 3,985 (14.8) |
|  | ≥ 50 and <60 | 6,332 (22.6) | 5,034 (18.7) |
|  | ≥ 60 and <70 | 5,386 (19.2) | 5,006 (18.6) |
|  | ≥ 70 | 3,896 (13.9) | 4,276 (15.9) |

### Sex and ancestry

We used the genetically inferred sex and ancestry estimated from the WES data by Regeneron in the analyses. Briefly, genetic ancestry was estimated using principal component analysis (PCA) of common SNPs shared with the HapMap3 reference panel^23^. PCs were computed using the reference samples, and study samples were projected onto them. Kernel density estimates of each reference population in PC1 vs PC2 and PC3 vs PC4 were used to assign each sample to matching reference populations, which were then mapped into five genetic ancestry groups: African (AFR), Admixed American (AMR), European (EUR), South Asian (SAS), and East Asian (EAS). Samples that matched no reference population, or an ambiguous combination of populations, were labeled Unknown.

### CHIP variant calling and classification

Somatic single-nucleotide variants (SNVs) and small insertions and deletions were called from aligned CRAM files using our previously described pipeline^24,25^, which implements GATK Mutect2 [4.2.0.0] in tumor-only mode with the GRCh38 reference genome. To exclude likely germline calls and sequencing artifacts, we provided to Mutect2 an external reference of germline variants from gnomAD and a Panel Of Normals (PON). For the gnomAD germline resource, we used the GATK-provided somatic-hg38_af-only-gnomad.hg38.vcf.gz resource. We generated separate PONs for the discovery and validation cohorts, each using WES data from 100 participants aged ≤25 years, an age group in which clonal hematopoiesis is expected to be uncommon. To minimize potential confounding from conditions associated with CHIP, the PON did not include individuals with neoplasms, HIV and inflammatory bowel disease (IBD). Finally, high-confidence somatic mutations were identified by applying Mutect2’s orientation bias and PASS filters.

Candidate variants were restricted to a curated set of myeloid CHIP-associated variants across 76 prespecified driver genes, and required to have a variant allele fraction (VAF) ≥2% (Supplementary Table S1). Additional filters to remove likely artifacts required a minimum read depth ≥20; ≥3 reads supporting the mutant allele; and at least one forward- and one reverse-strand read supporting both the reference and alternate alleles. Somatic variants were excluded if their gnomAD^26^ population allele frequency was ≥0.1% overall or in any ancestry-specific population. To reduce potential germline contamination, we filtered candidate CHIP variants using ancestry-specific allele frequencies rather than relying solely on overall population frequencies. This excluded variants such as *TET2* rs568009712, which was enriched among MSM participants of South Asian ancestry, showed elevated VAFs, and had an allele frequency of 0.4% in the South Asian population in gnomAD^26^. Variants with an observed allele frequency >1% in the cohort were also excluded, unless they had previously been implicated in hematologic malignancy. Variants were annotated for predicted pathogenicity using Combined Annotation Dependent Depletion (CADD) score (dbNSFP version 4.2a), and those with a Phred-scaled CADD score <10 were excluded. *U2AF1* required additional handling because the GRCh38 reference genome contains an incorrect duplication of the *U2AF1* gene locus on chromosome 21 that impairs read mapping. *U2AF1* CHIP variants were recovered using a custom *pileup region* script^8^. All CHIP variants that passed the filters are listed in Supplementary Table S2. Participants with at least one CHIP variant were classified as CHIP carriers; all other participants were classified as non-carriers. Clonal cytopenia of undetermined significance (CCUS) was not assessed separately.

### Clone size definitions

For each CHIP mutation, clone size was defined using mutation-level VAF. Mutations with VAF <10% were classified as small clones, and those with VAF ≥10% as large clones. Participant-level clone size analyses therefore use the maximum observed VAF for those individuals with multiple CHIP mutations.

### HLA typing and zygosity classification

To examine potential associations between HLA zygosity and CHIP, we inferred HLA class I and class II alleles for each participant from WES CRAM files using HLA-HD [1.7.1] at up to three-field (six-digit) resolution^27^. For zygosity analyses, allele calls were collapsed to two-field (four-digit) resolution. At each locus, participants carrying two distinct alleles were classified as heterozygous, whereas those carrying two identical alleles were classified as homozygous.

Participants heterozygous at all three classical HLA class I loci (*HLA-A*, *HLA-B*, *HLA-C*) were classified as fully class I heterozygous; participants homozygous at one or more loci were classified as non-fully class I heterozygous. Participants heterozygous at each of five classical HLA class II loci (*HLA-DRB1*, -*DQB1*, -*DQA1*, -*DPB1*, and -*DPA1*) were classified as fully class II heterozygous; participants homozygous at one or more loci were classified as non-fully class II heterozygous. We also evaluated locus-specific zygosity at *HLA-DRA* and the non-classical HLA class II loci (*HLA-DMA*, *HLA-DMB*, *HLA-DOA*, and *HLA-DOB*). Participants with missing or ambiguous calls (∼0-25% depending on the loci, Supplementary Table S3) were excluded from analyses of the corresponding locus; composite class I and class II analyses required complete calls at all constituent loci.

### Systemic immune-inflammation index

Given the reported association between inflammation and CHIP^28^, we considered two blood-based indices of systemic inflammation. The systemic Immune-Inflammation Index (SII) is a composite marker that integrates innate inflammatory responses and adaptive immune function, calculated from peripheral blood cell counts as platelet count × neutrophil count / lymphocyte count, with higher values indicating greater systemic inflammation. We also considered the INFLA-score as a complementary measure of low-grade systemic inflammation because it combines C-reactive protein (CRP) with cellular inflammatory markers^19^. However, CRP measurements were available for only a limited subset of participants. Analyses therefore focused on SII, which could be calculated for a larger proportion of participants from available blood cell counts.

Laboratory measurements were obtained from MSM health records and were subjected to data cleaning and harmonization, including unit standardization and removal of implausible, non-measurement, or sentinel values. The empirical distribution of each laboratory variable was subsequently examined to identify distributional gaps and extreme observations. These observations were evaluated alongside clinically plausible ranges to derive each measurement specific admissible value ranges: platelet count: 0-2,500 K/µL, neutrophils absolute count: 0-100 K/µL, lymphocyte absolute count: 0-300 K/µL. As part of the lab data quality control, we further excluded measurements that were recorded on at least three consecutive days as they could be indicative of acute clinical conditions or an inpatient monitoring. We also excluded multiple measurements that were available on the same day, which could reflect inpatient monitoring or repeat testing. For each participant, we selected the complete blood count obtained closest to the biospecimen collection date within a one-year window. To ensure that the SII was calculated from a single blood draw, all three components (platelet, neutrophil, and lymphocyte counts) were required to be available on the same date.

### Replication of previously reported germline variant associations with CHIP risk

In a recent study, Kessler et al.^29^ conducted common and rare variant associations in the UK Biobank cohort, comprising 25,657 CHIP cases and 342,869 controls of European ancestry. The study identified 57 independent common (minor allele frequency (MAF) > 0.5%) germline variants across 24 loci, as well as a rare germline frameshift variant in the *CHEK2* gene associated with the risk of acquiring CHIP. More recently, a multi-ancestry meta-analysis of six large-scale cohorts (UKB, All of Us Research Program, TOPMed, Vanderbilt University Biobank, Mass General Brigham Biobank, and Mexico City Prospective Study) including 53,274 CHIP cases and 922,528 controls identified 31 genome-wide significant loci harboring 2,572 variants^10^. We assessed whether these 2,593 previously reported germline variant associations with CHIP risk were replicated in our cohort.

### Disease phenotypes

To evaluate the association between CHIP and disease phenotypes, we mapped EHR diagnoses coded using ICD-9 and ICD-10 diagnosis codes to disease phenotypes previously associated with CHIP^9^. Specifically, we evaluated i) hematological neoplasms, including all hematological neoplasms, myeloid malignancies, myeloproliferative neoplasms, myelodysplastic syndromes, acute myeloid leukemias, and lymphoid leukemias; ii) solid tumors, including non-hematological malignant neoplasms and cancers of the breast, prostate, lung, colorectum, melanoma, kidney, head and neck, brain, esophagus, stomach, liver, and thyroid; and iii) other conditions previously reported to be associated with CHIP, including cardiovascular disease (myocardial infarction, stroke, ischemic stroke, coronary artery disease, heart failure, atrial fibrillation, and all cardiovascular diseases), type 2 diabetes mellitus, hypertension, obesity, rheumatoid arthritis, Alzheimer’s disease, Parkinson’s disease, chronic kidney disease, gout, chronic obstructive pulmonary disease (COPD), HIV, and inflammatory bowel disease (IBD). We evaluated disease associations using two frameworks: (i) prevalent diseases at baseline and (ii) incident diseases occurring after biospecimen collection. The ICD-9/10 codes used to define each disease phenotype are provided in Supplementary Table S4.

For prevalent disease analysis, participants were considered cases if the corresponding diagnosis code was recorded on at least two distinct encounter dates before or on the biospecimen collection date. For incident disease analysis, participants with prevalent disease at baseline were excluded. Incident cases were defined as participants with the corresponding diagnosis code recorded on at least two distinct encounter dates after biospecimen collection. In both analyses, participants without a qualifying diagnosis code for the corresponding disease before or after biospecimen collection were considered controls. We also performed additional stratified analyses of disease incidence to evaluate associations with specific CHIP subgroups, including high- and low-VAF (<10%) CHIP, mutations in the most frequently observed CHIP genes (*DNMT3A* and *TET2*), mutations in high-risk CHIP genes (*TP53, ASXL1, JAK2, U2AF1, SF3B1, SRSF2, IDH1* and *IDH2*), and CHIP among young (18-39 years) and older adults (≥40 years).

### EHR phenotype construction and phenome-wide association study (PheWAS)

To characterize the clinical phenome associated with CHIP in this hospital-based cohort, we performed a phenome-wide association study (PheWAS) using the *PheWAS* R package (version 0.99.6.1)^30^. Clinical diagnoses were derived from longitudinal EHR data using ICD-9 and ICD-10 diagnosis codes and mapped to ∼1,800 phecodes using the *createPhenotypes* function. While mapping, phecode exclusions and sex-specific restrictions were applied. The incidence phenotype definitions included diagnoses recorded after biospecimen collection without a corresponding diagnosis documented before the biospecimen collection date. A participant was classified as a case for a phenotype when the corresponding phecode was documented on at least two distinct encounter dates. Participants without evidence of the phenotype were considered as controls; those with a single qualifying encounter were omitted from the analysis of that phenotype. Sex-specific phecodes were evaluated only among participants of the applicable sex. Participants were stratified according to age at biospecimen collection as young adults (18-39 years) or older adults (≥40 years). This stratification was used to examine marked age-related differences in both CHIP prevalence and overall disease burden.

### Statistical analysis

#### General approach and demographic variables

Analyses were performed in R. Unless otherwise specified, models were fitted separately in the discovery and validation cohorts. Effect estimates are reported as odds ratios (ORs) with 95% confidence intervals (CIs) and two-sided *p-*values. In addition to overall CHIP, gene-specific analyses were performed for the four most prevalent CHIP driver genes: *DNMT3A, TET2, ASXL1,* and *PPM1D*. Age was modeled continuously in years; sex (male/female), smoking status (yes/no), and genetic ancestry were modeled categorically. Genetic ancestry was categorized into the three predominant genetic ancestry groups in this cohort, EUR, AFR, and AMR, in addition to Other, with EUR as the reference group. Associations of age, sex, smoking status, and genetic ancestry with CHIP were first evaluated using univariable logistic regression, with CHIP carrier status as the binary outcome (carrier/non-carrier). Covariates significantly associated with CHIP status in univariable analyses were then included in a multivariable logistic regression model, with CHIP status as the binary outcome. Statistical significance was defined as *p* ≤ 0.05.

#### Clone size analysis

To account for potential differences in clone size distributions across genetic ancestry groups, among CHIP carriers, the association between age and clone size was evaluated separately within each major genetic ancestry group (EUR, AFR, and AMR) and cohort, resulting in six independent analyses. Logistic regression models used clone size (VAF≥10% vs. <10%) as the binary outcome and age in years as the predictor. The six estimates were then combined using random-effects meta-analysis with restricted maximum likelihood (REML) estimation and the Hartung–Knapp adjustment.

#### HLA zygosity analysis

To evaluate associations between HLA zygosity and CHIP, we used separate logistic regression models for each HLA locus and for the composite class I and class II measures. Locus-specific models compared heterozygous and homozygous participants; composite models compared fully heterozygous with non-fully heterozygous participants. Overall CHIP status was the binary outcome (carrier vs. non-carrier), and models were adjusted for age, sex and genetic ancestry. We performed additional analyses using *DNMT3A*-mutant and *TET2*-mutant CHIP as separate outcomes, comparing carriers of each mutation with participants without CHIP. Cohort-specific estimates were combined using inverse-variance weighted fixed-effects meta-analysis using the *metafor* R package^31^. To assess whether HLA associations were consistent across sequencing platforms and ancestry groups, we calculated Cochran’s (Q) and (I^2^) to assess and quantify whether variation among effect estimates exceeds that expected by chance. An (I^2^) of 0% indicates no detectable heterogeneity, whereas higher values indicate progressively greater differences in effect estimates across groups. As a secondary analysis, estimates from the six cohort-by-ancestry strata were combined using random-effects meta-analysis to assess the consistency of associations across genetic ancestry groups.

#### Systemic immune-inflammation index analysis

The association between SII and CHIP was evaluated using logistic regression with CHIP carrier status as the binary outcome, and log10-transformed SII as the predictor. Models were adjusted for age, sex and genetic ancestry, and restricted to participants with SII data. Estimates were obtained separately for discovery and validation cohorts and combined by inverse-variance-weighted fixed-effect meta-analysis. Additional analyses were performed to evaluate associations between SII and CHIP by clone size and individual CHIP driver genes.

#### Germline variant association analysis

Associations between previously reported germline variants and CHIP risk were evaluated using logistic regression, with CHIP carrier status as the binary outcome and genotype dosage, defined as the number of effect alleles (0, 1, or 2), as the predictor under an additive genetic model. Models were adjusted for age, sex, and genetic ancestry. Analyses were performed separately in the discovery and validation cohorts.

#### Disease phenotype analyses

Associations between CHIP carrier status and disease phenotypes were evaluated separately for prevalent and incident disease. For prevalent disease, we used a logistic regression model with disease status as the outcome and CHIP carrier status as the primary predictor. For incident disease, we used a Cox proportional hazards regression model to estimate hazard ratios (HRs) and 95% CIs, with time from biospecimen collection to the first qualifying diagnosis as the time-to-event measure. All models were adjusted for age, sex, and genetic ancestry. For stratified analyses of incident disease, the same Cox proportional hazards framework was applied. Analyses were restricted to disease phenotypes with at least two CHIP carriers among individuals with the disease. Effect estimates were obtained separately in the discovery and validation cohorts and pooled using inverse-variance-weighted fixed-effect meta-analysis. To account for multiple testing, we also report *p*-values adjusted using the Benjamini–Hochberg procedure.

#### PheWAS

For each phecode, logistic regression models used phecode case-control status as the binary outcome and CHIP carrier status as the primary predictor, adjusting for age, sex, and genetic ancestry. Models were fitted separately in the discovery and validation cohorts and within each age stratum (18-39 years and ≥ 40 years), based on age at biospecimen collection. Only phecodes represented by at least 20 cases or 20 controls were tested, with an additional requirement of at least 5 cases among CHIP carriers. Within each age stratum, cohort-specific estimates for phecodes tested in both cohorts were combined by inverse-variance weighted fixed-effects meta-analysis using the *metafor* R package^31^. Prior to meta-analysis, we excluded phecodes for which the ratio of the cohort-specific variances (squared standard errors) exceeded 100 to limit the influence of unstable effect estimates. To account for multiple testing, statistical significance was determined using a Bonferroni-corrected threshold, which was applied separately within each age stratum across the meta-analyzed phecodes. Associations that did not meet the multiple-testing-corrected significance threshold but with nominal *p* ≤ 0.05 were considered exploratory.

## Results

### Study design overview

We characterized CHIP prevalence, mutational spectrum, and clone size (VAF) across age and genetic ancestry groups in the Mount Sinai Million (MSM) Health Discoveries Program cohort (Figure 1). Subset analyses focused on young adults (18–39 years), as well as CHIP subgroups defined by clone size and driver gene. We also evaluated inherited genetic correlates of CHIP, including HLA heterozygosity and previously reported common and rare germline variants, and assessed associations with blood-based inflammatory index. Finally, we integrated genomic data with longitudinal electronic health records (EHRs) to conduct age-stratified (18–39 years and ≥40 years) phenome-wide association studies (PheWAS) using clinical phenotypes derived from ICD-9 and ICD-10-codes.

**Figure 1.**
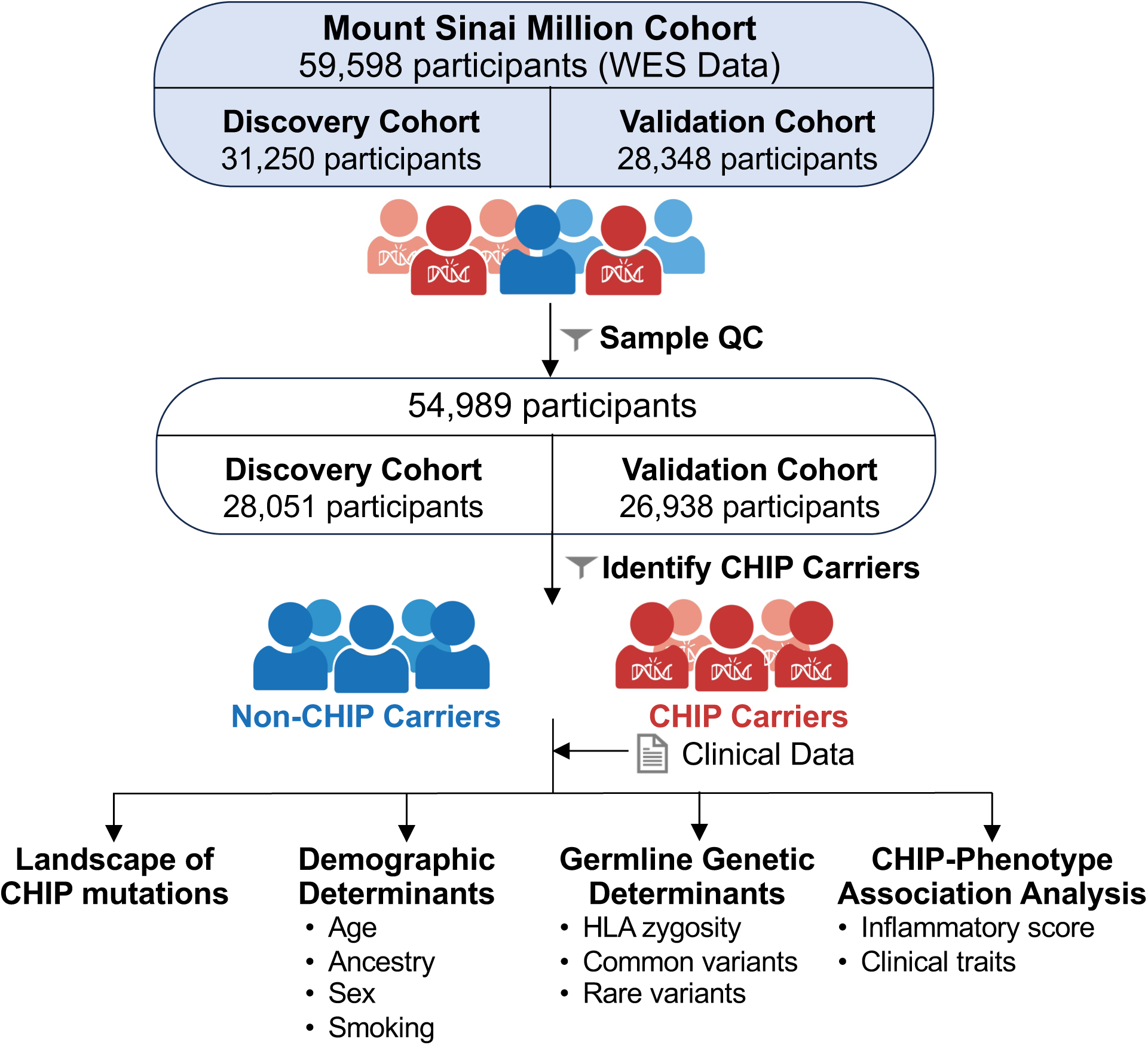
Study design and overview of CHIP characterization in the Mount Sinai Million cohort.

### Cohort characteristics

Of 59,598 MSM participants, 59,011 passed sample QC and 54,989 remained after subsequent exclusions. These participants were then divided into discovery (n = 28,051) and validation (n = 26,938) cohorts based on sequencing methodology (see Methods). Demographic characteristics are summarized in Table 1. Participants in the discovery cohort were aged 18-82 years (median: 52 years), with major genetic ancestry groups 36% European (EUR), 41% African (AFR), and 12% Admixed American (AMR). Participants in the validation cohort were aged 18-87 years (median: 51 years), and 38%, 28%, and 18% were assigned EUR, AFR, and AMR genetic ancestry, respectively. Across the combined study population, 37% were of EUR, 35% of AFR, and 15% of AMR genetic ancestry. Ancestry-stratified analyses focused on these three major groups.

### CHIP mutational landscape in MSM cohort

We identified 2,647 CHIP mutations among 2,386 participants (8.5%) in the discovery cohort, and 2,629 CHIP mutations among 2,284 individuals (8.5%) in the validation cohort. Across both cohorts and the three major ancestry groups, around 90% of CHIP carriers harbored a single mutation (Figure 2A-B).

**Figure 2.**
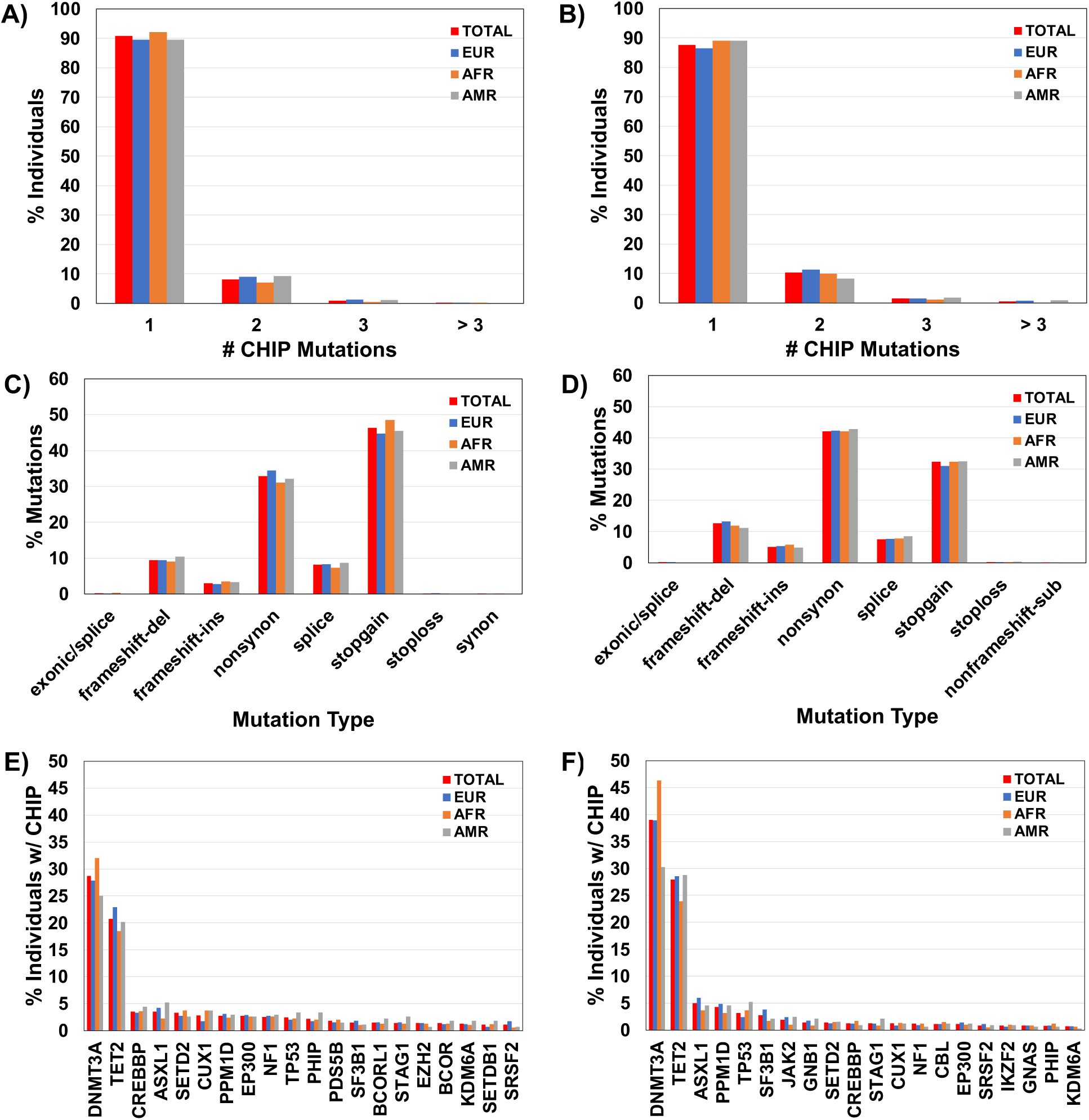
Characteristics of CHIP mutations in the discovery and validation cohorts stratified by genetic ancestry. Characteristics of CHIP mutations in the discovery (A, C, E) and validation cohorts (B, D, F), stratified by genetic ancestry (EUR, blue; AFR, orange; AMR, gray) and overall (TOTAL, red). **A-B)** Percentage of CHIP carriers with 1,2, 3 or >3 CHIP mutations**, C-D)** Percentage of CHIP mutations by mutation type **E-F)** Percentage of individuals with CHIP carrying mutations in the top 20 most frequently mutated genes

Nonsynonymous and stopgain mutations were the predominant mutation types (Figure 2C-D). The most frequently mutated genes were *DNMT3A*, *TET2*, *ASXL1* and *PPM1D* (Figure 2E-F), consistent with the established CHIP mutational landscape.

### Young adult (age 18-39 years) CHIP carriers in the MSM cohort

Unlike population-based biobanks, which tend to include predominantly middle-aged and older participants, the MSM cohort included 15,974 (∼30%) participants aged 18-39 years (7,337 in the discovery cohort; 8,637 in the validation cohort). Of these, 646 (4.0%) were CHIP carriers, including 383 (5.2%) in the discovery cohort and 263 (3.0%) in the validation cohort. The median age of CHIP carriers was 31 years in both cohorts. The majority of young adult CHIP carriers harbored a single mutation (Supplementary Figure S1). As in the overall cohort, nonsynonymous and stop-gain mutations were the predominant mutation types. The most frequently mutated genes were *TET2*, *DNMT3A*, and *CUX1*. Mutation characteristics and ancestry distributions among young adults are shown in Supplementary Figure S1. In comparison, older adults (age ≥40 years) had higher CHIP prevalence in both cohorts (9.7% vs. 5.2% in the discovery cohort; 11.0% vs. 3.0% in the validation cohort), with the top mutated genes broadly similar (Supplementary Table S5).

### CHIP prevalence increased with age and varied by genetic ancestry

CHIP prevalence increased with age in both cohorts and within each of the three major genetic ancestry groups (Figure 3). Prevalence also differed across ancestry groups. In the discovery cohort, CHIP was detected in 9.7% of EUR, 8.0% of AFR, and 8.2% of AMR participants; the corresponding prevalences in the validation cohort were 10.5%, 8.0%, and 6.6%, respectively. For young adults, CHIP prevalence among EUR, AFR, and AMR participants was 5.3%, 5.0%, and 6.0% in the discovery cohort, and 2.8%, 2.4% and 3.4%, in the validation cohort respectively.

**Figure 3.**
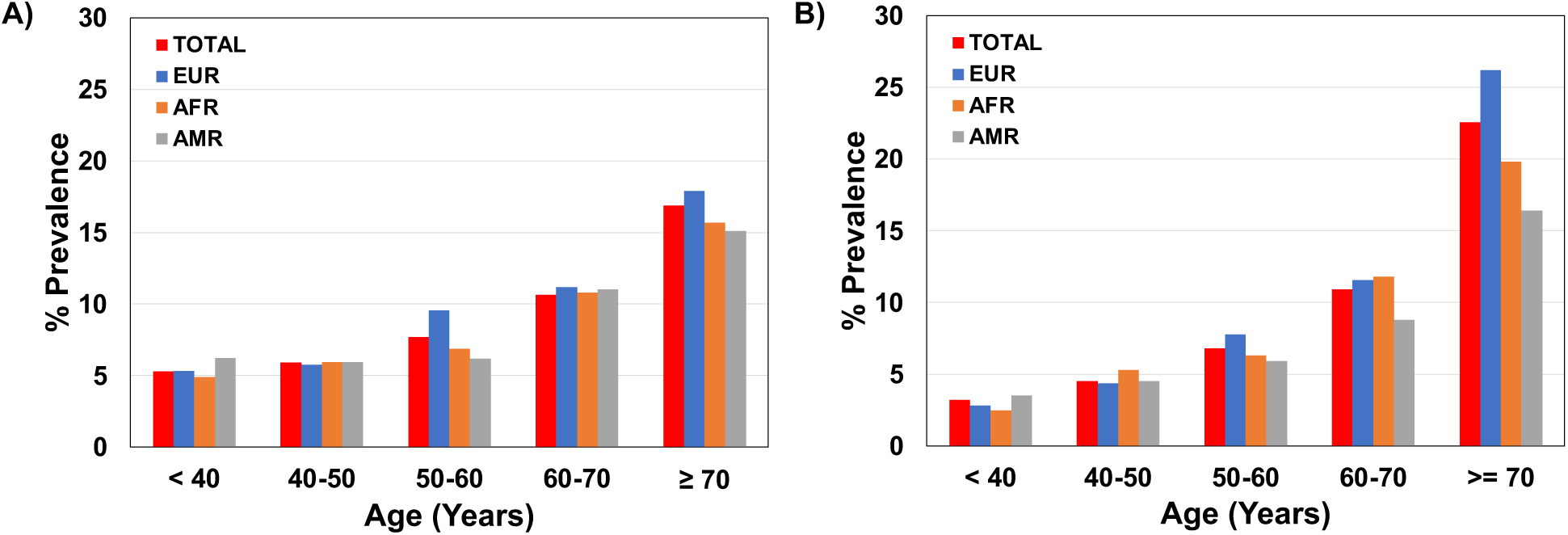
CHIP prevalence in the discovery and validation cohorts stratified by genetic ancestry. Percentage of CHIP carriers within each age group in the **A)** discovery **B)** and validation cohorts, stratified by genetic ancestry (EUR, blue; AFR, orange; AMR, gray) and overall (TOTAL, red).

We first evaluated the associations between individual demographic variables and CHIP using univariable regression (Table 2). CHIP was significantly associated with age and genetic ancestry, but not with sex or smoking status. In multivariable analyses adjusting for genetic ancestry, each additional year of age was associated with higher odds of CHIP in both cohorts (discovery: OR=1.030, 95% CI = 1.027–1.033, *p* <2e-16; validation: OR=1.051, 95% CI = 1.048–1.054, *p* <2e-16). After adjusting for age, AFR participants had lower odds of CHIP than EUR participants in both cohorts (discovery: OR=0.86, 95% CI = 0.78–0.95, *p*=0.002; validation: OR=0.84, 95% CI = 0.75–0.93, *p*=0.001). AMR ancestry was associated with lower estimated odds of CHIP, although the association did not reach statistical significance in the discovery cohort (discovery: OR=0.88, 95% CI = 0.76–1.01, *p*=0.08; validation: OR=0.72, 95% CI = 0.63– 0.82, *p*=8.47e-07). Complete results are provided in Table 2.

**Table 2.** Associations of demographic and clinical characteristics with CHIP status.

**A) Discovery cohort**
|  |  | CHIP carriers<br>(2,386) | CHIP non-carriers<br>(25,665) | Univariable regression | Multivariable regression |
| --- | --- | --- | --- | --- | --- |
|  |  | # Samples (%) | # Samples (%) | OR (p) [95% CI] | OR (p) [95% CI] |
| **Age (yrs)** | Median | 60 | 51 | 1.031 (<2e-16) [1.028-1.034] | 1.030 (<2e-16) [1.027-1.033] |
| **Sex** | Female | 1,403 (58.8) | 15,284 (59.6) | Ref |  |
|  | Male | 983 (41.1) | 10,381 (40.4) | 1.03 (0.48) [0.95 - 1.12] |  |
| **Smoking status** | No | 1,950 (81.7) | 20,750 (80.8) | Ref |  |
|  | Yes | 404 (16.9) | 4,550 (17.7) | 0.94 (0.32) [0.84 - 1.06] |  |
| **Genetic ancestry** | EUR | 969 (40.6) | 9,025 (35.2) | Ref |  |
|  | AFR | 916 (38.4) | 10,602 (41.3) | 0.80 (6.71e-6) [0.73 - 0.88] | 0.86 (0.002) [0.78 - 0.95] |
|  | AMR | 268 (11.2) | 2,999 (11.7) | 0.83 (0.01) [0.72 - 0.96] | 0.88 (0.08) [0.76 - 1.01] |
|  | Other | 233 (9.8) | 3,039 (11.8) | 0.71 (9.18e-6) [0.62 - 0.83] | 0.81 (0.007) [0.70 - 0.95] |

**B) Validation cohort**
|  |  | CHIP carriers<br>(2,284) | CHIP non-carriers<br>(24,654) | Univariable regression | Multivariable regression |
| --- | --- | --- | --- | --- | --- |
|  |  | # Samples (%) | # Samples (%) | OR (p) [95% CI] | OR (p) [95% CI] |
| **Age (yrs)** | Median | 67 | 50 | 1.052 (<2e-16) [1.049-1.055] | 1.051 (<2e-16) [1.048-1.054] |
| **Sex** | Female | 1,296 (56.7) | 14,409 (58.4) | Ref |  |
|  | Male | 988 (43.3) | 10,245 (41.6) | 1.07 (0.11) [0.98 - 1.17] |  |
| **Smoking status** | No | 982 (43.0) | 10,316 (41.8) | Ref |  |
|  | Yes | 286 (12.5) | 2,673 (10.8) | 1.12 (0.10) [0.98 - 1.29] |  |
| **Genetic ancestry** | EUR | 1,072 (46.9) | 9,168 (37.2) | Ref |  |
|  | AFR | 598 (26.2) | 6,896 (28.0) | 0.74 (2.27e-8) [0.67 - 0.82] | 0.84 (0.001) [0.75 - 0.93] |
|  | AMR | 327 (14.3) | 4,593 (18.6) | 0.61 (4.34e-14) [0.54 - 0.69] | 0.72 (8.47e-7) [0.63 - 0.82] |
|  | Other | 287 (12.6) | 3,997 (16.2) | 0.61 (1.72e-12) [0.54 - 0.70] | 0.83 (0.01) [0.72 - 0.96] |

At the driver gene level, focused analyses of the four most frequently mutated genes *DNMT3A, TET2, ASXL1*, and *PPM1D* revealed that their prevalence increased with age after adjusting for genetic ancestry in both cohorts (Supplementary Table S6, Supplementary Figure S2). Similar to overall CHIP, *TET2-* and *ASXL1-*mutant CHIP were less prevalent among AFR than EUR participants. Associations among AMR participants were directionally similar but did not consistently reach statistical significance across cohorts.

### Clone size showed age-associated patterns across cohorts and ancestry groups

We assessed clone size using VAF. The majority of CHIP mutations had a VAF below 10% (Figure 4A-B), but older participants generally had larger clones across both cohorts (Figure 4C-H). We then assessed the association between age and the presence of large clones (VAF≥10%) separately by cohort, modeling age per 1-year increase and adjusting for genetic ancestry and sex. Older age was associated with higher odds of a large clone in the discovery cohort (OR=1.04, 95% CI = 1.03–1.04, *p<*2e-16), with a consistent trend in the validation cohort (OR=1.01, 95% CI = 1.00–1.01, *p=*0.06). In driver gene level analyses (Supplementary Table S7), older age was associated with higher odds of large clones in *ASXL1* in both cohorts (discovery: OR=1.08, 95% CI = 1.03–1.13, *p*=0.003; validation: OR=1.04, 95% CI = 1.00–1.08, *p*=0.04). For *DNMT3A* and *TET2*, the associations were significant in the discovery cohort, and trended in the same direction in the validation cohort. *PPM1D* showed a similar trend but was not significantly associated with large clones in either cohort.

**Figure 4.**
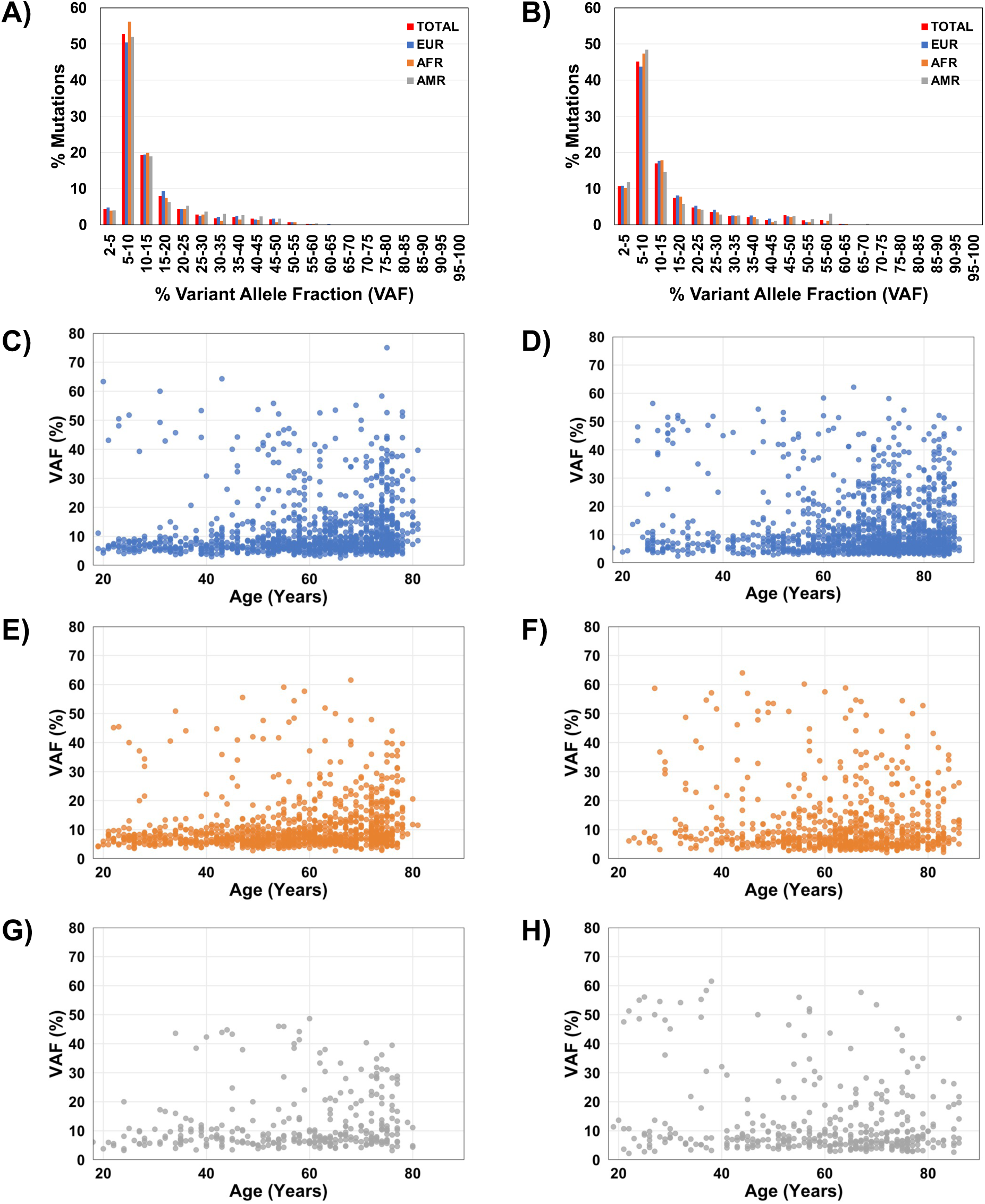
Landscape of chip variant allele fraction (VAF) distribution and age-related patterns across diverse ancestry groups. **A–B)** Distribution of CHIP mutations by VAF across total cohort and by genetic ancestry groups. Relationship between age and VAF for CHIP mutations stratified by ancestry groups C-D) EUR, E-F) AFR and G-H) AMR shown separately for the discovery and validation cohorts.

In a secondary analysis across cohort-by-ancestry strata, in the discovery cohort, increasing age was associated with higher odds of a large clone among EUR (OR=1.04, 95% CI=1.03– 1.05, *p<*2e-16), AFR (OR=1.04, 95% CI=1.03–1.05, *p*=8.54e-13) and AMR participants (OR=1.03, 95% CI=1.01–1.05, *p*=0.001). In the validation cohort, the association remained significant among EUR individuals (OR=1.01, 95% CI=1.00–1.02, *p*=0.002), but did not reach statistical significance among AFR or AMR participants (Supplementary Table S7). A random-effects meta-analysis of the cohort- and ancestry-specific estimates yielded a pooled OR of 1.02 (95% CI = 1.00–1.04, *p*=0.05) with substantial heterogeneity (I^2^ = 91%, p<0.001).

### Inherited immune variation and systemic inflammation in CHIP

We examined inherited HLA variation and systemic inflammation as immune-related correlates of CHIP. We assessed HLA variation through allele heterozygosity, and systemic inflammation through the systemic immune-inflammation index (SII). Analyses were performed separately in the discovery and validation cohorts, adjusting for age, sex and ancestry, and cohort-specific estimates were combined using fixed-effect meta-analysis.

### Certain HLA heterozygosities showed locus-, clone-size and driver gene-specific associations with CHIP

To better understand the potential immunogenomic drivers of CHIP, we evaluated associations between HLA allele heterozygosity and CHIP prevalence, adjusting for age, sex and ancestry separately in the discovery and validation cohorts, and combined cohort-specific estimates using fixed-effect meta-analysis (Figure 5). The proportions of individuals heterozygous at each HLA locus are provided in Supplementary Table S3. At a nominal (*p*≤0.05) threshold, *HLA-DPA1* heterozygosity was associated with 7% lower odds of overall CHIP (pooled OR=0.93, 95% CI = 0.87–0.99, *p*=0.03; Figure 5A). Point estimates indicated higher odds for *HLA-DQA1* heterozygosity (pooled OR=1.10, 95% CI = 1.00–1.22, *p*=0.05), and lower odds for *HLA-DQB1* heterozygosity (pooled OR=0.92, 95% CI = 0.84–1.02, *p*=0.10). No HLA class I locus or aggregate class I or class II heterozygosity measure was associated with overall CHIP at the *p* ≤ 0.05 threshold.

**Figure 5.**
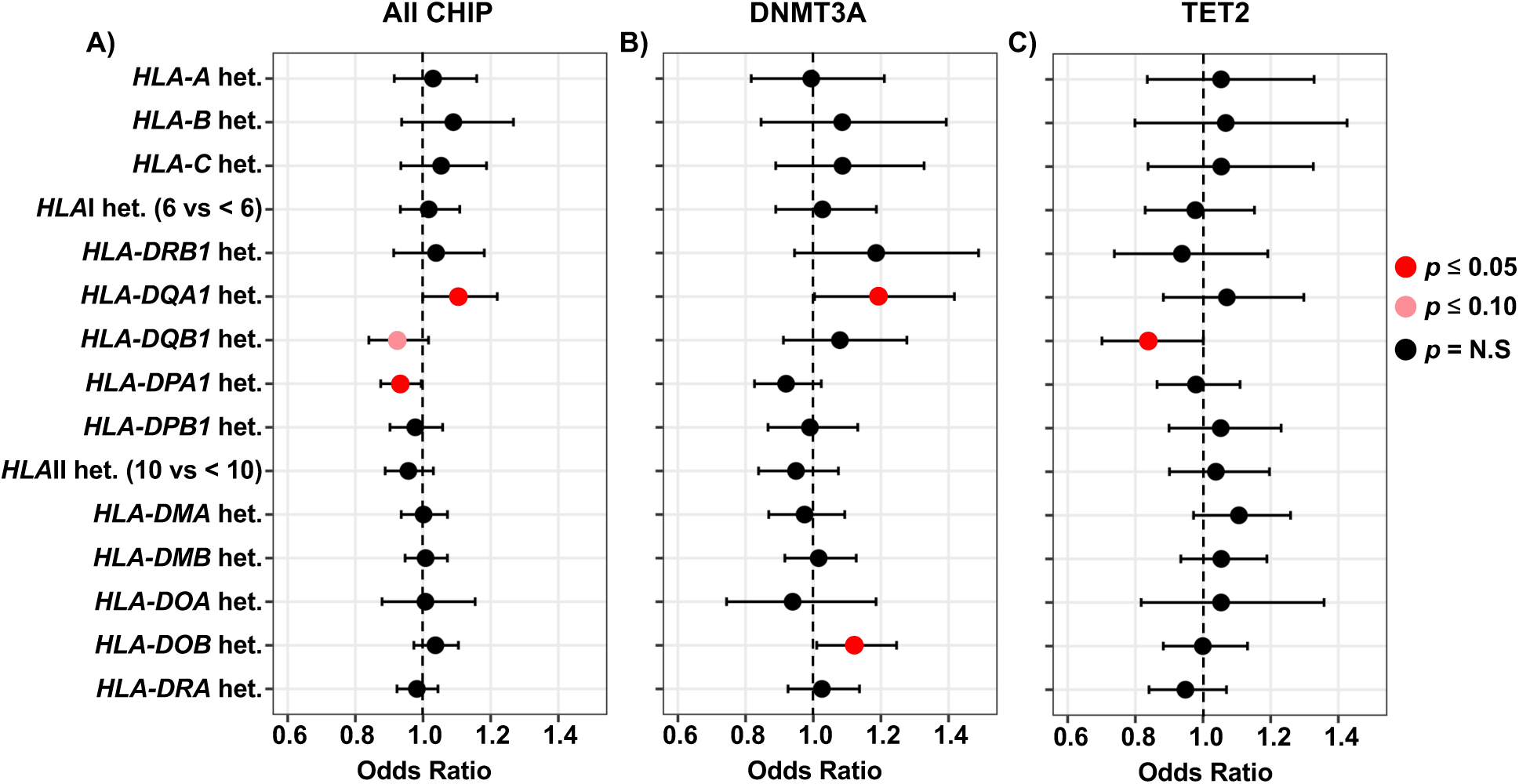
Association between HLA heterozygosity and CHIP prevalence. Forest plots showing odds ratios for the association between HLA heterozygosity and **A)** overall CHIP, **B)** *DNMT3A*-mutant CHIP, and **C)** *TET2*-mutant CHIP. Points represent the estimated ORs, and whiskers indicate the corresponding 95% confidence intervals. Results shown are from meta analyses of the discovery and validation cohorts. Color indicates significance level: red, *p* ≤ 0.05; pink, *p* ≤ 0.10; black, not significant (N.S.)

Among CHIP carriers, we additionally performed an exploratory analysis comparing maximum VAF ≥20% versus <20% to examine HLA associations with larger clones. A 20% threshold has been used in myeloid malignancy risk stratification, including the clonal hematopoiesis risk score^32^. We observed that HLA-C heterozygosity was nominally associated with higher odds of a maximum VAF ≥20% versus <20% (pooled OR=1.52, 95% CI = 1.19–1.94, *p*=0.001).

#### Driver gene-specific analyses revealed distinct HLA associations for DNMT3A- and TET2-mutant CHIP

Heterozygosity at both *HLA-DQA1* (pooled OR=1.19, 95% CI = 1.00–1.42, *p*=0.05) and *HLA-DOB* (pooled OR=1.12, 95% CI = 1.01–1.25, *p*=0.03) was associated with higher odds of *DNMT3A*-mutant CHIP (Figure 5B). *DNMT3A* R882 hotspot mutations were identified in 64 participants in the discovery cohort and 109 in the validation cohort. Excluding these carriers yielded similar effect estimates for *HLA-DQA1* heterozygosity (pooled OR=1.19, 95% CI=0.99–1.42, p=0.07) and *HLA-DOB* heterozygosity (pooled OR=1.12, 95% CI=1.00– 1.25, p=0.050), while R882-specific estimates were positive but imprecise. In contrast, *HLA-DQB1* heterozygosity associated with lower odds of *TET2*-mutant CHIP (pooled OR=0.84, 95% CI = 0.70–1.00, *p*=0.05; Figure 5C). Note that while *HLA-DQB1* effect estimates were opposite in direction in the *DNMT3A*- and *TET2*-mutant CHIP analyses, only the *TET2* estimates reached the statistical threshold (Supplementary Table S8). Collectively, these findings suggest that associations between HLA class II zygosity and CHIP show different patterns according to the underlying driver mutation.

#### Ancestry-stratified HLA analyses

To examine the HLA associations within genetic ancestry groups, we performed separate analyses among EUR, AFR and AMR participants in each cohort. Within each ancestry group, we combined the discovery and validation estimates using fixed-effect meta-analysis to obtain ancestry-specific pooled ORs. Among participants of European genetic ancestry, *HLA-DQA1* heterozygosity showed higher estimated odds of overall CHIP (pooled OR=1.15, 95% CI = 0.98–1.36, *p*=0.08) and was associated with higher odds of *DNMT3A-*mutant CHIP (pooled OR=1.49, 95% CI = 1.11–2.01, *p*=0.008). Among participants of African genetic ancestry, *HLA-DMA* heterozygosity was associated with higher odds of *TET2*-mutant CHIP (pooled OR=1.49, 95% CI = 1.16–1.91, *p*=0.002), with concordant estimates in both cohorts. A comparable association was not observed in the AMR ancestry groups. Additional ancestry-stratified and less common driver-specific analyses are summarized in Supplementary Table S8.

#### Secondary HLA meta-analysis across cohort-by-ancestry strata

We then combined all six cohort-by-ancestry estimates: EUR, AFR, and AMR within the discovery and validation cohorts, to obtain a second overall estimate and assess between-stratum heterogeneity. This analysis complemented the primary ancestry-adjusted analysis by pooling associations estimated separately within ancestry groups. For *DNMT3A*-specific *HLA-DQA1* heterozygosity, the pooled OR was 1.21 (95% CI = 0.97–1.51, *p*=0.074), with minimal estimated between-stratum heterogeneity (I^2^=0.54%, Q-test *p* = 0.52). Estimates for *HLA-DOB* heterozygosity and *DNMT3A*-mutant CHIP were directionally consistent across ancestry groups (pooled OR=1.14, 95% CI = 1.04–1.25, *p*=0.02; I^2^=0%; no detectable between-stratum heterogeneity). Estimates for *HLA-DQB1* heterozygosity and *TET2*-mutant CHIP were also directionally consistent across cohorts and ancestry groups (pooled OR=0.84, 95% CI = 0.71–1.00, *p*=0.06). Collectively, these findings identify locus-specific HLA associations with overall CHIP, clone size, and driver-defined CHIP subtypes and suggest that these associations may also vary across genetic ancestry groups.

### CHIP and systemic immune-inflammation index (SII)

We next examined systemic inflammation as a phenotypic correlate of CHIP. Eligible laboratory measurements for SII calculation were available for 67.6% of participants in the discovery cohort and 57.0% in the validation cohort. Higher log10-transformed SII was associated with increased odds of CHIP in both the discovery (OR=1.25, 95% CI = 1.07–1.48, *p*=0.005) and validation (OR=1.39, 95% CI = 1.12–1.71, *p*=0.002) cohorts (Figure 6, Supplementary Table S9). The pooled OR was 1.30 (95% CI =1.15–1.48, p=4.78e-5).

**Figure 6.**
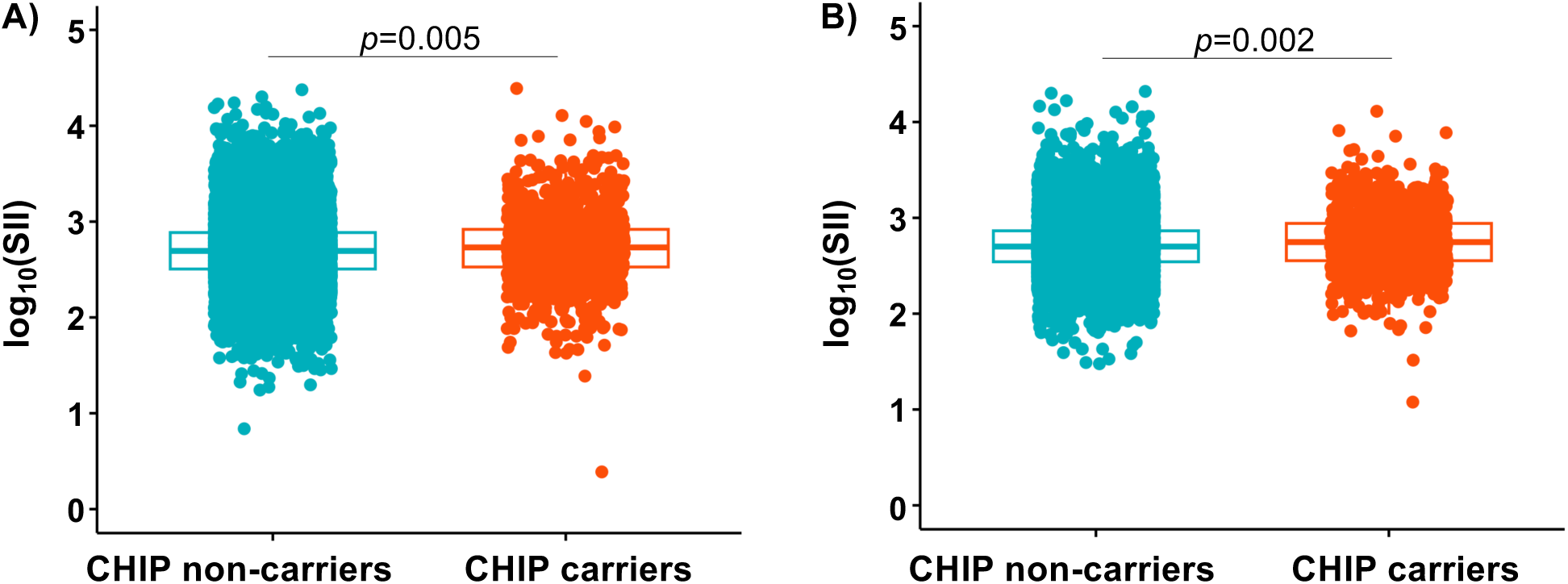
Systemic immune-inflammation index (SII) distribution based on CHIP status. Distribution of SII values in CHIP non-carriers (blue) versus CHIP carriers (red) in the **A)** discovery cohort and **B)** validation cohort. Each point represents an individual participant; boxplots indicate the median and interquartile range.

In gene-specific analyses, higher SII was associated with increased odds of *TET2*-mutant CHIP (pooled OR=1.40, 95% CI = 1.09–1.79, *p*=0.01) and *PPM1D*-mutant CHIP (pooled OR=2.33, 95% CI = 1.25–4.33, *p*=0.01). Age-stratified analyses showed similar associations among younger adults aged 18–39 years (pooled OR=1.58, 95% CI = 1.11–2.24, *p*=0.01) and older adults aged ≥40 years (pooled OR=1.21, 95% CI = 1.05–1.39, *p*=0.01). Associations were also observed for both low-VAF CHIP (pooled OR=1.25, 95% CI = 1.06–1.46, *p*=0.01) and high-VAF CHIP (pooled OR=1.35, 95% CI = 1.11–1.65, *p*=0.002).

### Inherited genetic determinants of CHIP

Of the 2,593 common variants previously identified^10,29^ as associated with CHIP risk in GWAS analyses, 2,474 were present in our cohort. Overall, 74 variants (Supplementary Table S10) were nominally significant in both cohorts including the *TERT* variant (rs7705526-A, 5:1285859:C:A) (discovery: OR=1.09, 95% CI = 1.02–1.16, p=0.01; validation: OR=1.25, 95% CI = 1.17–1.34, p=8.66e-11), that had the strongest association from the previous study^10^.

### Associations between CHIP and prevalent and incident disease outcomes vary by clone size and underlying driver genes

We evaluated associations between CHIP and prevalent and incident outcomes across three categories: hematologic neoplasms, solid tumors, and other conditions previously linked to CHIP, including cardiovascular disease. We estimated ORs for prevalent disease (Supplementary Table S11) and hazard ratios (HRs) for incident disease (Figure 7, Supplementary Table S11).

**Figure 7.**
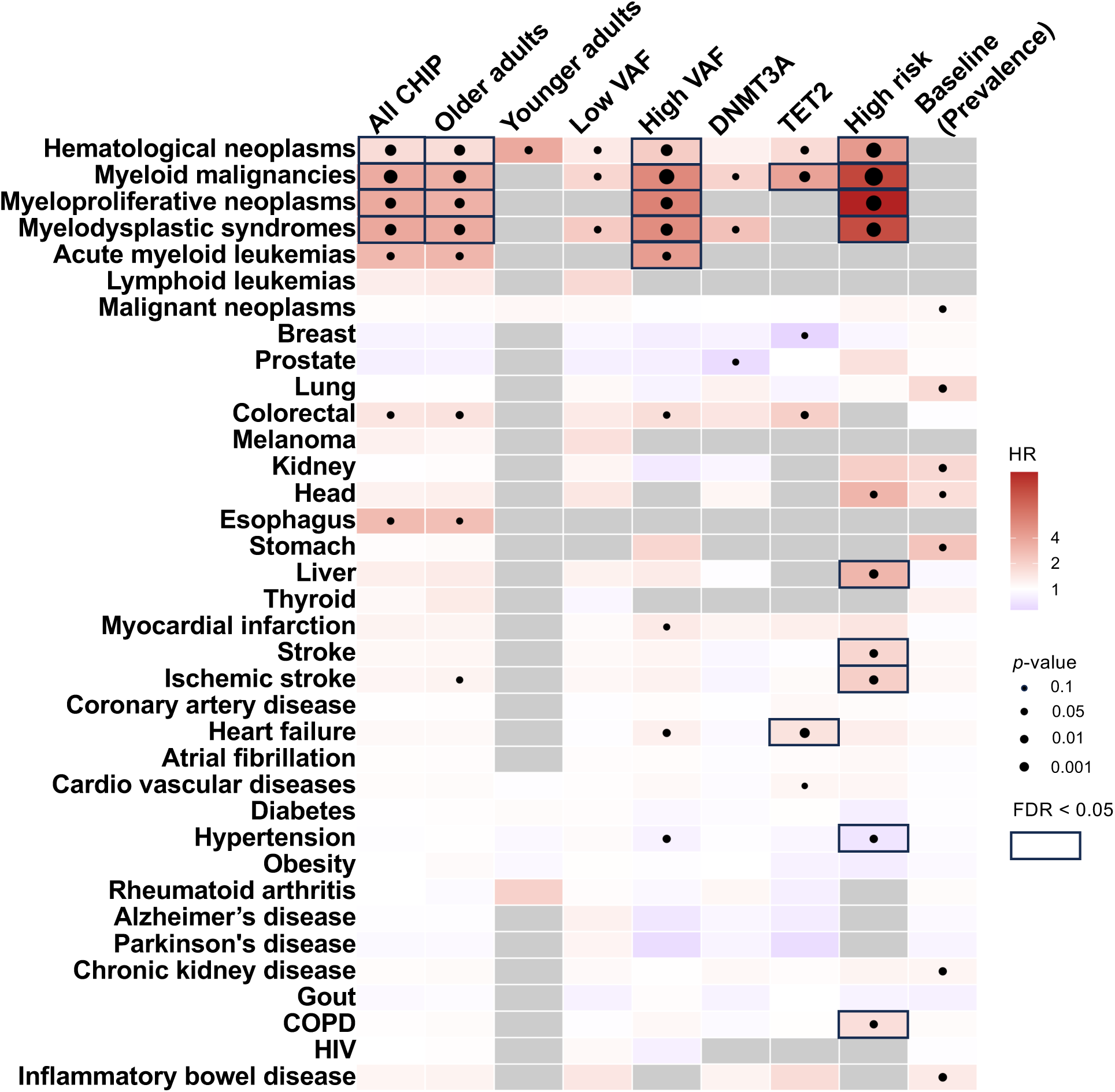
Associations between CHIP and incident diseases. Heatmaps showing associations with overall CHIP; CHIP among older and younger adults; CHIP with large (VAF ≥10%) and small (VAF < 0.1) clones; and CHIP driven by *DNMT3A, TET2*, and high-risk genes. The red–blue color scale represents the hazard ratio (HR). Gray indicates missing HR estimates; black dots indicate associations *p* ≤ 0.1; and black squares indicate associations that remained significant after false discovery rate (FDR) correction (FDR *p* ≤ 0.05).

#### Hematologic neoplasms

CHIP was associated with higher incidence of myeloid malignancies (pooled HR=3.54, 95% CI = 2.46–5.09, *p*=1.16e-11), myeloproliferative neoplasms (pooled HR=3.64, 95% CI = 2.06–6.44, *p*=8.97e-6), and myelodysplastic syndromes (pooled HR=3.70, 95% CI = 2.20–6.23, *p*=8.54e-07). These associations were generally higher for CHIP with large clones, including myeloid malignancies (pooled HR=5.97, 95% CI = 3.91–9.12, *p<*2e-16); myeloproliferative neoplasms, (pooled HR=6.61, 95% CI = 3.41–12.82, *p*=2.19e-8); and myelodysplastic syndromes (pooled HR=5.63, 95% CI = 3.06–10.35, *p*=2.71e-08). The associations were highest for CHIP involving high-risk genes: myeloid malignancies (pooled HR=15.71, 95% CI = 9.59–25.74, *p<*2e-16), myeloproliferative neoplasms (pooled HR=24.16, 95% CI =11.50–50.74, *p<*2e-16), and myelodysplastic syndromes (pooled HR=14.44, 95% CI = 7.23–28.84, *p*=4.01e-14). Among young adults, CHIP showed a nominal association with incident hematologic malignancies overall. At gene level, *TET2*-mutant CHIP was significantly associated with myeloid malignancies (pooled HR=4.06, 95% CI = 2.21–7.45, *p*=6.45e-6).

#### Solid tumors

For solid tumors, we generally observed positive associations across several incident and prevalent analyses for colorectal, liver, esophageal, lung, stomach, head and neck and kidney cancers. The association between CHIP involving high-risk genes and incident liver cancer remained statistically significant after FDR correction (pooled HR=3.00, 95% CI = 1.52 – 5.92, *p*=0.002), whereas the other solid-tumor associations did not.

#### Cardiovascular and other diseases

Cardiovascular associations were observed primarily for CHIP with large clones (VAF ≥10%), *TET2*-mutant CHIP, and CHIP involving high-risk genes. Particularly, *TET2*-mutant CHIP was associated with heart failure. CHIP involving high-risk genes was associated with higher HRs of ischemic stroke (pooled HR=2.05, 95% CI = 1.30– 3.24, *p*=0.002), stroke (pooled HR = 1.86, 95% CI = 1.18–2.93, *p*=0.007), and COPD (pooled HR=1.63, 95% CI = 1.09–2.44, *p*=0.01) and a lower hazard of hypertension (pooled HR=0.70, 95% CI = 0.53–0.93, *p*=0.02). Stratification by clone size suggested that associations were predominantly driven by high-VAF clones (VAF≥10%) rather than low-VAF clones. Complete results by clone size and driver gene are provided in Supplementary Table S11.

### CHIP exhibited age-stratified associations with clinical phenotypes in PheWAS

To characterize clinical associations more broadly, we performed PheWAS separately among young adults aged 18-39 years (discovery cohort: n=7,337; validation cohort: n=8,637) and adults aged ≥40 years (discovery: n=20,714; validation: n=18,301). Analyses included incident diagnoses documented after biospecimen collection for sequencing. Associations were estimated separately in the discovery and validation cohorts and combined by fixed-effect meta-analysis (Figure 8, Supplementary Table S12).

**Figure 8.**
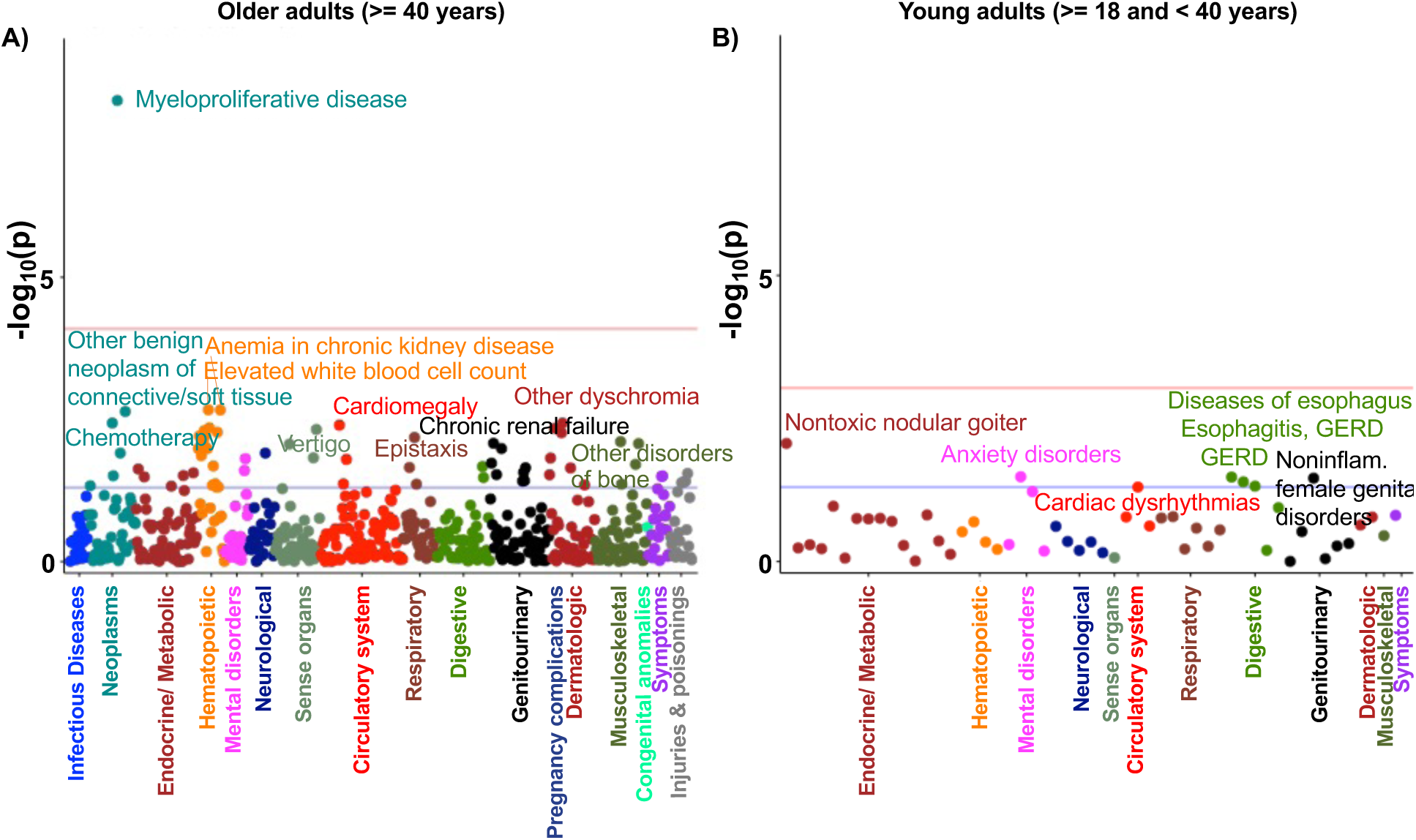
Phenome-wide associations between CHIP and disease phenotypes stratified by age group. Manhattan plots showing the results of the PheWAS meta-analysis across the discovery and validation cohorts for **A)** older adults (age ≥40 years) **B)** young adults (18 ≤ age <40 years). Each point represents a phecode tested for association with CHIP carrier status. The blue horizontal line indicates the significance threshold *p* = 0.05 and the red horizontal line indicates the phenome-wide significance threshold after correction for multiple testing.

Among adults aged ≥40 years, 622 phecodes were included in the meta-analysis. The strongest association was with myeloproliferative disease (phecode 200; OR=3.32, 95% CI = 2.21-5.00, *p*=7.79e-9), which remained significant after Bonferroni correction. Additional nominal associations were observed across hematopoietic and neoplastic phenotypes, including diseases of white blood cells (phecode 288; OR=1.48, 95% CI = 1.12–1.94, *p*=0.005), pancytopenia (phecode 284.1; OR=1.82, 95% CI = 1.20–2.75, *p*=0.005), other anemias (phecode 285; OR=1.20, 95% CI = 1.05–1.36, *p*=0.006), and iron deficiency anemia (phecode 280; OR=1.23, 95% CI = 1.05–1.45, *p*=0.010). Nominal associations were also observed with benign neoplasms of connective and other soft tissue (phecode 215; OR=1.72, 95% CI = 1.21– 2.44, *p*=0.002), chemotherapy (phecode 197; OR=1.29, 95% CI = 1.09–1.53, *p*=0.004), and leukemia (phecode 204; OR=1.56, 95% CI = 1.10–2.21, *p*=0.012).

Outside these categories, nominal associations included cardiomegaly (OR=1.73, 95% CI = 1.19–2.50, *p*=0.004), chronic renal failure (OR=1.28, 95% CI = 1.07–1.53, *p*=0.008), and disorders resulting from impaired renal function (OR=1.72, 95% CI = 1.14–2.61, *p*=0.010). Additional associations were observed with dermatologic conditions, including dyschromia and vitiligo (OR=0.73, 95% CI = 0.59–0.91, *p*=0.005), as well as disorders of the sense organs, including light-headedness and vertigo (OR=1.21, 95% CI = 1.06–1.37, *p*=0.005) and hypermetropia (OR=1.85, 95% CI = 1.17–2.92, *p*=0.009).

Among young adults, only 54 phecodes met the eligibility thresholds for minimum number of participants in both cohorts (see Methods). The strongest observed association was with nontoxic nodular goiter (phecode 241; OR=2.02, 95% CI = 1.20–3.41, *p*=0.009). Additional associations included several digestive phenotypes, particularly diseases of esophagus (phecode: 530, OR=1.42, 95% CI = 1.03–1.97, *p*=0.03) including esophagitis, gastroesophageal reflux disease (GERD), and related diseases. CHIP was also negatively associated with anxiety disorders (phecode: 300, OR=0.67, 95% CI = 0.46–0.97, *p*=0.03). Other associations included noninflammatory female genital disorders (OR = 1.42, 95% CI = 1.03–1.97, *p* = 0.03) and cardiac dysrhythmias (OR = 1.39, 95% CI = 1.00–1.93, *p* = 0.050) (Supplementary Table S12). Together, these findings identify clinical phenotypes associated with CHIP within each age stratum.

Among older adults, associations were predominantly hematologic and neoplastic phenotypes, with additional associations involving genitourinary and dermatologic disorders. In contrast, among younger adults, the nominal associations were predominantly nonhematologic. Given the limited number of phecodes that could be evaluated in younger adults, we also examined phenotypes that showed suggestive associations (*p* ≤ 0.05) in both age groups. Nontoxic nodular goiter was the only overlapping nominal association in both age groups, captured under related phecodes (young adults: phecode 241; OR = 2.02, 95% CI = 1.20–3.41, *p* = 0.009, older adults: phecode 241.2; OR = 1.41, 95% CI = 1.05–1.89, *p* = 0.02). Complete association results are provided in Supplementary Table S12.

## Discussion

In this study, we characterized CHIP in adults from the Mount Sinai Million Health Discoveries Program (MSM), a multi-ancestry, EHR-linked hospital cohort spanning adulthood. Substantial representation of African and Admixed American genetic ancestry and adults younger than 40 years enabled examination of CHIP in populations less well-represented in large sequencing studies. Linking genomic data with longitudinal EHRs, we examined the CHIP mutational landscape alongside inherited HLA variation, systemic inflammation, and clinical outcomes. Associations with lower CHIP prevalence among participants of African ancestry, and higher systemic immune-inflammatory index (SII) were observed in both discovery and validation cohorts. Together, these analyses extend the demographic, immune, and clinical characterization of CHIP across adulthood.

CHIP prevalence increased with age in both discovery and validation cohorts. After adjusting for age, participants of African (AFR) ancestry exhibited lower odds of CHIP than participants of European ancestry in both cohorts, and those of Admixed American (AMR) ancestry trended similarly. The latter pattern is consistent with findings from the Mexico City Prospective Study, in which clonal hematopoiesis prevalence was lower than in the UK Biobank, and increased with the proportion of European ancestry within the Mexican cohort^33^. Our findings extend these population-based observations to a clinically ascertained hospital cohort and highlight the importance of ancestry representation when characterizing CHIP prevalence.

Uniquely, the MSM cohort included 15,974 adults aged 18–39 years, an age group that is underrepresented in population biobanks. Their inclusion provided an opportunity to characterize CHIP in early adulthood. CHIP was detected in 646 young adults, corresponding to a combined prevalence of 4.0%. Most young carriers harbored a single mutation, with *TET2, DNMT3A,* and *CUX1* among the most frequently mutated genes. Young-adult prevalence differed between the discovery and validation cohorts, at 5.2% and 3.0%, respectively, compared with ∼1-3% observed in other cohorts^8,10^. Our results suggest that young-adults with CHIP are a subgroup that requires future longitudinal studies of clone persistence, growth, and clinical consequences.

We then asked whether HLA-mediated immunosurveillance impacts CHIP. A recent UK Biobank study found no consistent association between predicted MHC binding of recurrent CH-derived peptides and CH prevalence or clone size, and no protective association of greater HLA diversity^14^. However, that study focused on 40 recurrent variants in 11 genes, excluding *TET2*, and its heterozygosity analysis evaluated classical HLA loci in participants of European genetic ancestry. Here, we asked whether these findings extend to a broader mutation spectrum and to nonclassical class II loci involved in peptide editing. Towards this end, we examined HLA zygosity across classical and nonclassical loci in the multi-ancestry MSM cohort, evaluating associations with overall CHIP, clone size, and driver-defined subtypes.

Our exploratory analyses identified nominal associations involving several HLA class II loci. *HLA-DPA1* heterozygosity was associated with lower odds of overall CHIP, whereas *HLA-DQA1* and *HLA-DOB* heterozygosity was associated with higher odds of *DNMT3A*-mutant CHIP. These loci encompass both peptide-presenting molecules and regulators of peptide editing. HLA class II molecules are expressed primarily by antigen-presenting cells, where HLA-DQ and HLA-DP present peptides to CD4 T-cells, while HLA-DM facilitates peptide exchange, and HLA-DO regulates HLA-DM activity, helping shape repertoire of presented peptides. These findings span both classical peptide-presenting molecules and nonclassical regulators of peptide editing^34–36^.

As the HLA genotype is inherited, it predates CHIP and can play a role in determining which peptides derived from an acquired mutant clone can be presented to T-cells. The observed differences in the direction of some HLA class II effect estimates between *DNMT3A*- and *TET2*-mutant CHIP suggest that HLA variation may drive the selection of clones carrying these mutations. Mutations in *DNMT3A* and *TET2* can generate distinct but partly overlapping epigenetic and transcriptional states^37–39^: *DNMT3A* catalyzes *de novo* DNA methylation, whereas *TET2* oxidizes 5-methylcytosine as part of the demethylation pathway. These states could alter antigen-processing pathways or the repertoire of available peptides, potentially affecting how mutant clones are recognized within a given HLA background.

A recent experimental study in AML has shown that the *DNMT3A*-R882H mutation generates an HLA-DQ–restricted neoantigen recognized by a CD4 T-cell clone through specific DQA1/DQB1 combinations^40^. In this study, HLA-DOB heterozygosity was associated with higher odds of *DNMT3A*-mutant CHIP in both primary and secondary meta-analyses. The similar effect estimates after excluding R882 mutations suggest that the observed *DNMT3A*–HLA associations may extend beyond this hotspot, motivating investigation of immune recognition across a broader *DNMT3A* mutation spectrum. Together with our *HLA-DQA1* finding, these observations support further investigation of HLA-DQ presentation and class II peptide editing in the recognition and persistence of *DNMT3A*-mutant clones. Allele- and haplotype-level analyses could clarify whether these associations reflect zygosity itself or particular HLA backgrounds. The exploratory association between *HLA-DMA* heterozygosity and *TET2*-mutant CHIP among participants of African genetic ancestry also motivates studying whether these relationships differ across HLA allele and haplotype backgrounds.

Among CHIP carriers, *HLA-C* heterozygosity was nominally associated with higher odds of a maximum VAF≥20% versus <20%. *HLA-C* is a class I molecule that participates in both adaptive and innate immune recognition: it presents peptides to CD8+ T-cells and serves as a ligand for killer-cell immunoglobulin-like receptors (KIRs), which regulate natural killer (NK) cell activity^41–43^. These dual functions suggest examining both T-cell and NK-cell pathways in relation to clone size. Longitudinal studies incorporating HLA-C alleles and KIR genotypes could help determine whether these immune interactions relate to subsequent clone persistence or expansion.

Independent replication and comparisons between driver-defined groups in larger cohorts will be needed to establish the specificity of these exploratory patterns. Allele- and haplotype-level analyses could clarify whether the associations reflect heterozygosity itself or particular HLA backgrounds, while functional studies could test their consequences for antigen presentation and immune recognition.

We also observed higher systemic immune-inflammation index (SII) values associated with CHIP in both cohorts. Because SII is derived from neutrophil, lymphocyte, and platelet counts, it may reflect both systemic inflammation and changes in hematopoietic output. Experimental studies, particularly in models of *TET2*-deficient hematopoiesis, support a bidirectional relationship between CHIP and inflammation: CHIP-associated mutations can promote inflammatory signaling, while inflammatory stress can favor the survival and expansion of mutant clones^15,16^. Although our findings are consistent with this relationship, they cannot distinguish whether inflammation precedes clonal expansion, results from altered hematopoiesis, or reflects shared clinical factors. SII may also be influenced by acute illness, medication use, and comorbid conditions^18,44,45^. When blood counts and DNA samples are collected at different times, SII may not reflect the inflammatory state at CHIP assessment. Repeated blood counts and sequencing would help clarify how changes in systemic inflammation relate to clone acquisition and expansion over time.

The targeted disease analyses and age-stratified PheWAS identified the clearest clinical associations with hematologic disease. Incident myeloid malignancies, myeloproliferative neoplasms, and myelodysplastic syndromes were associated with CHIP, with higher estimated hazard ratios in analyses of large clones and high-risk driver genes. Myeloproliferative disease also emerged as the strongest PheWAS association among adults aged ≥40 years and remained significant after correction for multiple testing. These findings emphasize the importance of considering clone size and driver mutations when examining the clinical correlates of CHIP. Although participants with hematologic malignancies diagnosed before or within six months after biospecimen collection were excluded, occult or prodromal disease at sampling cannot be fully ruled out.

Associations beyond hematologic disease were more limited after correcting for multiple testing. The targeted analyses nevertheless identified cardiovascular associations in selected CHIP subgroups, including nominal stroke associations involving high-risk genes (*TP53, ASXL1, JAK2, U2AF1, SF3B1, SRSF2, IDH1* and *IDH2*). Thus, the absence of cardiovascular phenotypes among the leading PheWAS findings should be considered alongside the targeted analyses. Differences in phenotype definitions, statistical models, and multiple-testing burden may contribute to the different patterns observed across these analyses. Similarly, nominal associations with anemia and renal phenotypes may reflect several processes, including hematopoietic changes, comorbid disease, and clinical ascertainment. The associations with chronic kidney disease are consistent with emerging human and experimental evidence linking CHIP to kidney-function decline, impaired recovery from acute kidney injury, and renal inflammation and fibrosis^46,47^.

The young-adult PheWAS findings were exploratory. Only 54 phecodes met the eligibility criteria in both cohorts, compared with 622 among adults aged ≥40 years, limiting direct comparison of the clinical patterns across age groups. The nominal associations reported in younger adults provide candidates for further investigation. Evaluating shared phenotypes in larger young-adult populations, with tests of differences by age, would help distinguish age-related variation from differences in statistical power and disease frequency.

Several limitations should be considered. WES at the sequencing depths used here has limited sensitivity for small clones, and differences in capture methods and sequencing coverage may affect comparisons between our discovery and validation cohorts despite using the same variant calling pipeline and thresholds. Tumor-only variant calling may retain residual germline variants or technical artifacts despite population-frequency filters and cohort-specific panels of normals. A single sequencing time point precludes direct measurement of clone growth. HLA alleles were inferred from WES, and locus-level heterozygosity cannot distinguish effects attributable to specific alleles, class II α–β pairings, or linked haplotypes. Testing multiple HLA loci, driver-specific outcomes, VAF thresholds, and ancestry strata creates a substantial multiple-testing burden. Associations supported only at nominal significance should therefore be considered exploratory and require independent replication. Broad genetic ancestry categories may obscure within-group diversity, and incomplete smoking information limits adjustment for this potential confounder. SII was available only for a subset of participants, which may introduce selection related to laboratory testing. It may also reflect hematopoietic output and concurrent clinical conditions in addition to inflammation. Statistical power was lower in young adults, limiting interpretation of age-stratified findings. Finally, hospital-based recruitment and EHR-derived phenotypes are subject to selection, encounter-frequency, and diagnostic misclassification biases. Diagnoses recorded after biospecimen collection establish the order of recorded events but may not identify the biological onset of disease, and the diagnostic exclusion window may not fully eliminate occult hematologic disease at sampling.

Overall, this study extends the characterization of CHIP across adulthood, and establishes MSM as a resource for studying CHIP in a multi-ancestry hospital population, including younger adults. The findings reproduce the established relationships of CHIP prevalence and clone size with age, identify ancestry-associated differences in prevalence, and characterize inflammatory and clinical correlates of CHIP. Exploratory HLA associations suggest that inherited HLA variation and systemic inflammation represent complementary components of the host immune context associated with CHIP. The locus-, clone size-, driver-, and ancestry-dependent HLA patterns are consistent with a model in which immune selection contributes to CHIP heterogeneity rather than exerting a uniform effect across clones. Longitudinal clone tracking, HLA haplotype analyses across ancestry groups, functional antigen-presentation studies, and targeted clinical outcome analyses will clarify the biological relevance of these associations and their implications for clonal evolution and disease risk.

## Data Availability

Mount Sinai Million data from Mount Sinai Health system requires PPHS/IRB approval.

## Acknowledgements

Research reported in this publication was supported by the Tisch Cancer Center at the Icahn School of Medicine at Mount Sinai, the Clinical and Translational Science Awards (CTSA) grant (KL2TR004421) from the Icahn School of Medicine at Mount Sinai, and in part through the Minerva computational and data resources and staff expertise provided by Scientific Computing and Data at the Icahn School of Medicine at Mount Sinai.

## Data availability

Mount Sinai Million WES data from Mount Sinai Health system requires PPHS/IRB approval.

## Code availability

All analyses were performed utilizing standard publicly available software. Any specific analysis code details are available from the authors upon request.

---

**Supplementary Figure S1.**
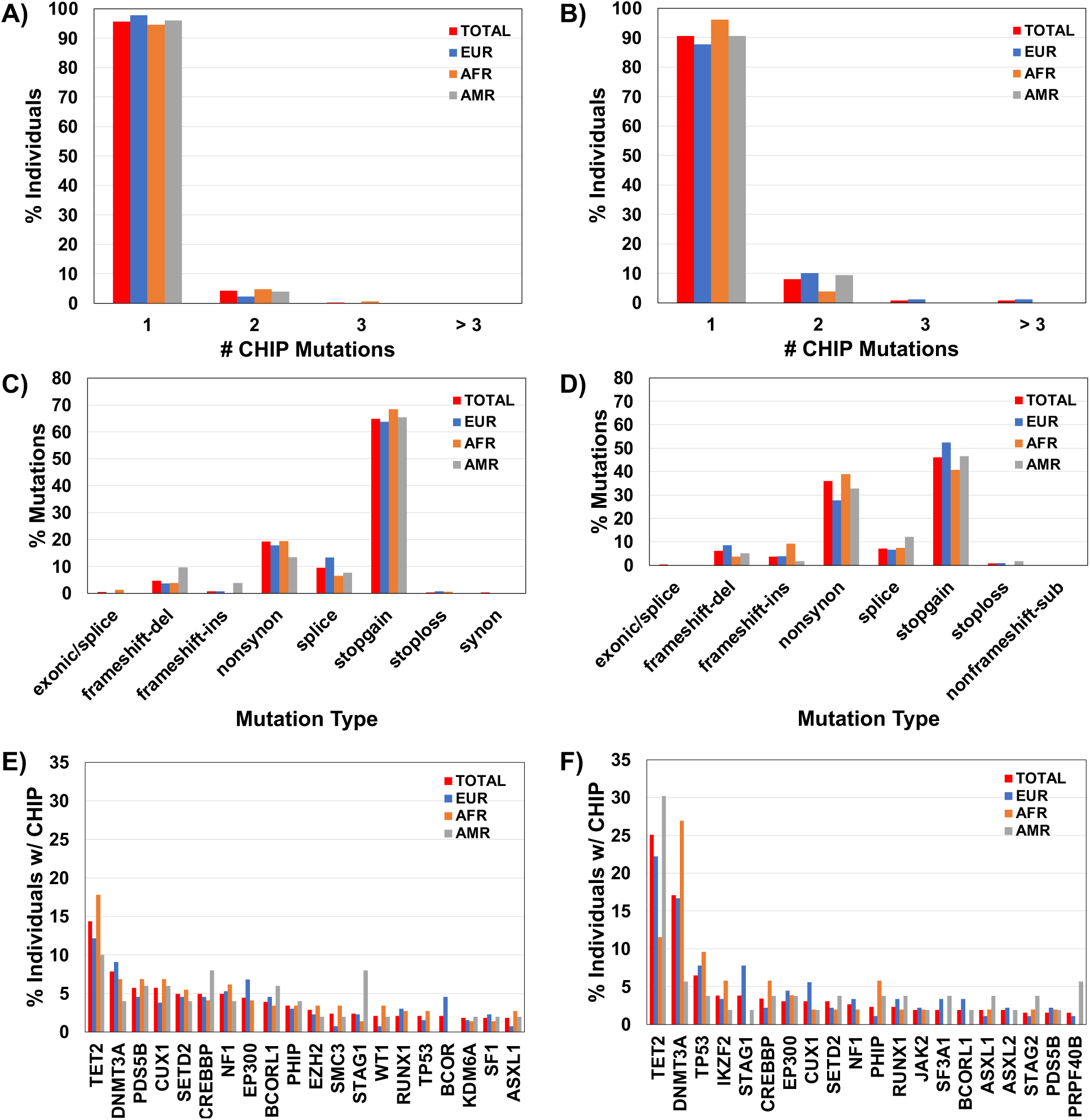
Characteristics of CHIP mutations among the young adults in the discovery and validation cohorts stratified by genetic ancestry. Characteristics of CHIP mutations in the discovery (A, C, E) and validation cohorts (B, D, F), stratified by genetic ancestry (EUR, blue; AFR, orange; AMR, gray) and overall (TOTAL, red). A-B) Percentage of CHIP carriers with 1,2, 3 or >3 CHIP mutations, C-D) Percentage of CHIP mutations by mutation type E-F) Percentage of individuals with CHIP carrying mutations in the top 20 most frequently mutated genes

**Supplementary Figure S2.**
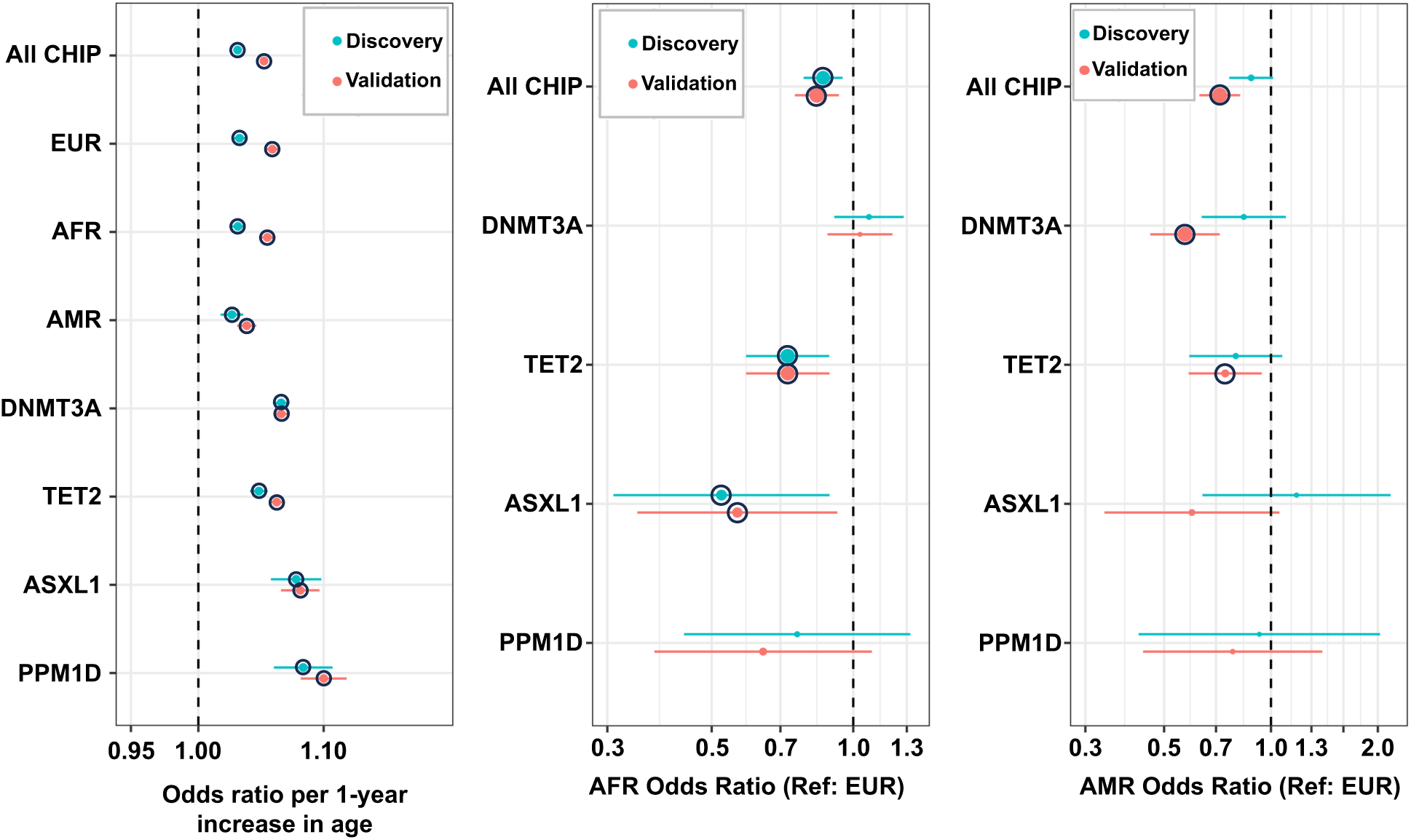
Associations of CHIP and individual CHIP genes with age and genetic ancestry across discovery and validation cohorts. A) Odds ratios (ORs) for CHIP prevalence associated with a 1-year increase in age. B) ORs for CHIP prevalence associated with African (AFR) versus European (EUR) genetic ancestry. C) ORs for CHIP prevalence associated with Admixed American (AMR) versus European (EUR) genetic ancestry. Points represent odds ratios, and whiskers indicate 95% confidence intervals. Associations were estimated using multivariable logistic regression models adjusting for age and genetic ancestry. The dashed vertical line indicates the null value (OR = 1). Black circles indicate associations with *p* ≤ 0.05.

## Supplementary Tables

**Supplementary Table S1.** List of myeloid associated CHIP mutations considered in this study

**Supplementary Table S2.** List of CHIP mutations identified among the **A)** 28,051 participants in the discovery cohort **B)** 26,938 participants in the validation cohort

**Supplementary Table S3.** Distribution of HLA gene zygosity in the discovery and validation cohorts

**Supplementary Table S4.** ICD-9/ICD-10 codes used to define different disease phenotypes

**Supplementary Table S5.** Comparison of the prevalence and top mutated genes in young adults (age <40 years) vs older adults (age ≥40 years) in the discovery and validation cohorts

**Supplementary Table S6.** Associations of demographic and clinical characteristics with CHIP by driver gene

**Supplementary Table S7.** Association of age with large-clone (VAF ≥10%) CHIP by driver gene and genetic ancestry

**Supplementary Table S8.** Association between HLA heterozygosity and CHIP prevalence by driver gene, VAF ≥20% and by genetic ancestry

**Supplementary Table S9.** Association between systemic immune-inflammation index and CHIP carrier status

**Supplementary Table S10.** Previously reported CHIP-associated GWAS variants nominally significant (p ≤ 0.05) in both the discovery and validation cohorts

**Supplementary Table S11.** Association between CHIP and prevalent and incident disease outcomes by age, clone size and driver gene

**Supplementary Table S12.** Top phenome-wide associations between CHIP and disease phenotypes among **A)** older adults (age ≥40 years) from meta-analysis **B)** young adults (age <40 years) from meta-analysis

## Notes

### Competing Interest Statement

The authors have declared no competing interest.

### Author Declarations

The Program for the Protection of Human Subjects (PPHS) at the Icahn School of Medicine at Mount Sinai determined that this study met the criteria for exempt human-subjects research (IRB no. 20-03271)

## References

1. Genovese G, Kähler AK, Handsaker RE, et al. Clonal hematopoiesis and blood-cancer risk inferred from blood DNA sequence. N Engl J Med. 2014;371(26):2477–2487. doi:10.1056/NEJMOA1409405

2. Khoury JD, Solary E, Abla O, et al. The 5th edition of the World Health Organization Classification of Haematolymphoid Tumours: Myeloid and Histiocytic/Dendritic Neoplasms. Leukemia. 2022;36(7):1703–1719. doi:10.1038/S41375-022-01613-1

3. Jaiswal S, Ebert BL. Clonal hematopoiesis in human aging and disease. Science. 2019;366(6465). doi:10.1126/SCIENCE.AAN4673

4. Jaiswal S, Fontanillas P, Flannick J, et al. Age-Related Clonal Hematopoiesis Associated with Adverse Outcomes. New England Journal of Medicine. 2014;371(26):2488–2498. doi:10.1056/NEJMOA1408617/SUPPL_FILE/NEJMOA1408617_DISCLOSURES.PDF

5. Heuser M, Thol F, Ganser A. M E D I C I N E Clonal Hematopoiesis of Indeterminate Potential A Risk Factor for Hematologic Neoplasms. Dtsch Arztebl Int. 2016;113(18):317–322. doi:10.3238/arztebl.2016.0317

6. Jaiswal S, Natarajan P, Silver AJ, et al. Clonal Hematopoiesis and Risk of Atherosclerotic Cardiovascular Disease. N Engl J Med. 2017;377(2):111–121. doi:10.1056/NEJMOA1701719

7. Bick AG, Weinstock JS, Nandakumar SK, et al. Inherited causes of clonal haematopoiesis in 97,691 whole genomes. Nature 2020 586:7831. 2020;586(7831):763-768. doi:10.1038/s41586-020-2819-2

8. Vlasschaert C, Mack T, Heimlich JB, et al. A practical approach to curate clonal hematopoiesis of indeterminate potential in human genetic data sets. Blood. 2023;141(18). doi:10.1182/BLOOD.2022018825

9. Kar SP, Quiros PM, Gu M, et al. Genome-wide analyses of 200,453 individuals yield new insights into the causes and consequences of clonal hematopoiesis. Nat Genet. 2022;54(8):1155–1166. doi:10.1038/S41588-022-01121-Z

10. Uddin MM, Yu Z, Weinstock JS, et al. Genomic and phenomic landscape of clonal hematopoiesis in over a million ancestrally diverse participants. medRxiv. 2026;27:2022.07.29.22278015. doi:10.1101/2022.07.29.22278015

11. Liu J, Tran D, Xue L, et al. Germline genetic variation impacts clonal hematopoiesis landscape and progression to malignancy. Nat Genet. 2025;57(8):1872–1880. doi:10.1038/S41588-025-02250-X;SUBJMETA

12. Chowell D, Morris LGT, Grigg CM, et al. Patient HLA class I genotype influences cancer response to checkpoint blockade immunotherapy. Science. 2018;359(6375):582–587. doi:10.1126/SCIENCE.AAO4572

13. Krishna C, Tervi A, Saffern M, et al. An immunogenetic basis for lung cancer risk. Science. 2024;383(6685):1–18. doi:10.1126/SCIENCE.ADI3808

14. Walkowiak B, MacGregor HAJ, Blundell JR. No evidence of immunosurveillance in mutation-hotspot-driven clonal hematopoiesis. Nat Genet. 2026;58(7):1643–1650. doi:10.1038/S41588-026-02594-Y

15. Fuster JJ, MacLauchlan S, Zuriaga MA, et al. Clonal hematopoiesis associated with TET2 deficiency accelerates atherosclerosis development in mice. Science. 2017;355(6327):842–847. doi:10.1126/SCIENCE.AAG1381

16. Cai Z, Kotzin JJ, Ramdas B, et al. Inhibition of Inflammatory Signaling in Tet2 Mutant Preleukemic Cells Mitigates Stress-Induced Abnormalities and Clonal Hematopoiesis. Cell Stem Cell. 2018;23(6):833–849.e5. doi:10.1016/J.STEM.2018.10.013

17. Yu Z, Fidler TP, Ruan Y, et al. Genetic modification of inflammation- and clonal hematopoiesis-associated cardiovascular risk. J Clin Invest. 2023;133(18). doi:10.1172/JCI168597

18. Hu B, Yang XR, Xu Y, et al. Systemic immune-inflammation index predicts prognosis of patients after curative resection for hepatocellular carcinoma. Clin Cancer Res. 2014;20(23):6212–6222. doi:10.1158/1078-0432.CCR-14-0442

19. Bonaccio M, Di Castelnuovo A, Pounis G, et al. A score of low-grade inflammation and risk of mortality: prospective findings from the Moli-sani study. Haematologica. 2016;101(11):1434–1441. doi:10.3324/HAEMATOL.2016.144055

20. Shen X, Song Y, Zhao H, et al. Routine inflammatory indices modify clonal hematopoiesis-related prognostic risk in patients undergoing percutaneous coronary intervention: a real-world cohort study. BMC Med. 2026;24(1). doi:10.1186/S12916-026-04711-4

21. Belbin GM, Cullina S, Wenric S, et al. Toward a fine-scale population health monitoring system. Cell. 2021;184(8):2068–2083.e11. doi:10.1016/j.cell.2021.03.034

22. Abul-Husn NS, Soper ER, Odgis JA, et al. Exome sequencing reveals a high prevalence of BRCA1 and BRCA2 founder variants in a diverse population-based biobank. Genome Med. 2019;12(1). doi:10.1186/s13073-019-0691-1

23. Altshuler DM, Gibbs RA, Peltonen L, et al. Integrating common and rare genetic variation in diverse human populations. Nature. 2010;467(7311):52–58. doi:10.1038/NATURE09298

24. Esai Selvan M, Nathan DI, Guisado D, et al. Clonal Hematopoiesis of Indeterminate Potential in Crohn’s Disease and Ulcerative Colitis. Inflamm Bowel Dis. Published online January 6, 2025. doi:10.1093/IBD/IZAE312

25. Selvan ME, Kuan PF, Yang | Xiaohua, et al. Distinct Characteristics of Lymphoid and Myeloid Clonal Hematopoiesis in World Trade Center First Responders. Am J Hematol. 2025;0:1–5. doi:10.1002/AJH.70014

26. Chen S, Francioli LC, Goodrich JK, et al. A genome-wide mutational constraint map quantified from variation in 76,156 human genomes. bioRxiv. Published online October 10, 2022:2022.03.20.485034. doi:10.1101/2022.03.20.485034

27. Kawaguchi S, Higasa K, Shimizu M, Yamada R, Matsuda F. HLA-HD: An accurate HLA typing algorithm for next-generation sequencing data. Hum Mutat. 2017;38(7):788–797. doi:10.1002/HUMU.23230

28. Hajishengallis G, Chavakis T. Inflammageing and clonal haematopoiesis interplay and their impact on human disease. Nat Rev Mol Cell Biol. 2026;27(5):377–393. doi:10.1038/S41580-025-00936-Y

29. Kessler MD, Damask A, O’keeffe S, et al. Common and rare variant associations with clonal haematopoiesis phenotypes. Nature |. 2022;612. doi:10.1038/s41586-022-05448-9

30. Carroll RJ, Bastarache L, Denny JC. R PheWAS: data analysis and plotting tools for phenome-wide association studies in the R environment. Bioinformatics. 2014;30(16):2375–2376. doi:10.1093/BIOINFORMATICS/BTU197

31. Viechtbauer W. Conducting Meta-Analyses in R with the metafor Package. J Stat Softw. 2010;36(3):1–48. doi:10.18637/JSS.V036.I03

32. Weeks LD, Niroula A, Neuberg D, et al. Prediction of risk for myeloid malignancy in clonal hematopoiesis. NEJM evidence. 2023;2(5). doi:10.1056/EVIDOA2200310

33. Wen S, Kuri-Morales P, Hu F, et al. Comparative analysis of the Mexico City Prospective Study and the UK Biobank identifies ancestry-specific effects on clonal hematopoiesis. Nat Genet. 2025;57(3):572–582. doi:10.1038/S41588-025-02085-6

34. Weber DA, Evavold BD, Jensen PE. Enhanced dissociation of HLA-DR-bound peptides in the presence of HLA-DM. Science. 1996;274(5287):618–620. doi:10.1126/SCIENCE.274.5287.618

35. Pos W, Sethi DK, Wucherpfennig KW. Mechanisms of peptide repertoire selection by HLA-DM. Trends Immunol. 2013;34(10):495–501. doi:10.1016/J.IT.2013.06.002

36. Perraudeau M, Taylor PR, Stauss HJ, et al. Altered major histocompatibility complex class II peptide loading in H2-O-deficient mice. Eur J Immunol. 2000;30(10):2871–2880. doi:10.1002/1521-4141(200010)30:10<2871::aid-immu2871>3.0.co;2-b

37. Nam AS, Dusaj N, Izzo F, et al. Single-cell multi-omics of human clonal hematopoiesis reveals that DNMT3A R882 mutations perturb early progenitor states through selective hypomethylation. Nat Genet. 2022;54(10):1514–1526. doi:10.1038/S41588-022-01179-9

38. Rasmussen KD, Jia G, Johansen J V., et al. Loss of TET2 in hematopoietic cells leads to DNA hypermethylation of active enhancers and induction of leukemogenesis. Genes Dev. 2015;29(9):910–922. doi:10.1101/GAD.260174.115

39. Zhang X, Su J, Jeong M, et al. DNMT3A and TET2 compete and cooperate to repress lineage-specific transcription factors in hematopoietic stem cells. Nat Genet. 2016;48(9):1014–1023. doi:10.1038/NG.3610

40. van der Lee DI, Argiro EM, Laan SNJ, et al. Mutated DNMT3A creates a public HLADQ-binding neoantigen on acute myeloid leukemia. Front Immunol. 2025;16. doi:10.3389/FIMMU.2025.1556121

41. Rasmussen M, Harndahl M, Stryhn A, et al. Uncovering the peptide-binding specificities of HLA-C: a general strategy to determine the specificity of any MHC class I molecule. J Immunol. 2014;193(10):4790–4802. doi:10.4049/JIMMUNOL.1401689

42. Older Aguilar AM, Guethlein LA, Adams EJ, Abi-Rached L, Moesta AK, Parham P. Coevolution of killer cell Ig-like receptors with HLA-C to become the major variable regulators of human NK cells. J Immunol. 2010;185(7):4238–4251. doi:10.4049/JIMMUNOL.1001494

43. Faure M, Barber DF, Takahashi SM, Jin T, Long EO. Spontaneous clustering and tyrosine phosphorylation of NK cell inhibitory receptor induced by ligand binding. J Immunol. 2003;170(12):6107–6114. doi:10.4049/JIMMUNOL.170.12.6107

44. Fois AG, Paliogiannis P, Scano V, et al. The Systemic Inflammation Index on Admission Predicts In-Hospital Mortality in COVID-19 Patients. Molecules. 2020;25(23). doi:10.3390/MOLECULES25235725

45. Satis S. New Inflammatory Marker Associated with Disease Activity in Rheumatoid Arthritis: The Systemic Immune-Inflammation Index. Curr Health Sci J. 2021;47(4):553–557. doi:10.12865/CHSJ.47.04.11

46. Kestenbaum B, Bick AG, Vlasschaert C, et al. Clonal Hematopoiesis of Indeterminate Potential and Kidney Function Decline in the General Population. Am J Kidney Dis. 2023;81(3):329–335. doi:10.1053/J.AJKD.2022.08.014

47. Vlasschaert C, Robinson-Cohen C, Chen J, et al. Clonal hematopoiesis of indeterminate potential is associated with acute kidney injury. Nat Med. 2024;30(3):810–817. doi:10.1038/S41591-024-02854-6

